## Supplementary material for "Occurrence and transmission potential of asymptomatic and presymptomatic SARS-CoV-2 infections: update of a living systematic review and meta-analysis"

| File name | Title | Pg # |
|---|---|---|
| **Checklists** | | |
| **S1 PRISMA Checklist** | **PRISMA Checklist 2020 and PRISMA Checklist for abstracts** | **2** |
| **Text** | | |
| **S1 Text** | **Search strings** | **6** |
| **S2 Text** | **Risk of Bias tool** | **8** |
| **Tables** | | |
| **S1 Table** | **Studies included in version 3.0 and excluded in version 4.0 of the living systematic review** | **13** |
| **S2 Table** | **Characteristics of studies reporting on proportion of asymptomatic SARS-CoV-2 infections (review question 1)** | **16** |
| **S3 Table** | **Location of studies contributing data to review question 1** | **38** |
| **S4 Table** | **Subgroup analysis according to risk of bias** | **40** |
| **S5 Table** | **Characteristics of mathematical modelling studies** | **41** |
| **Figures** | | |
| **S1 Fig** | **Flowchart of identified, excluded, and included records as of 31 January 2020** | **49** |
| **S2 Fig** | **Forest plot of proportion of people with asymptomatic SARS-CoV-2 infection, stratified by study design.** | **50** |
| **S3 Fig** | **Risk of bias assessment of studies in question 1 and 2.1** | **51** |
| **S4 Fig** | **Forest plot of proportion of people with asymptomatic SARS-CoV-2 infection by date of publication.** | **57** |
| **S5 Fig** | **Assessment of credibility of mathematical modelling studies.** | **58** |
| **Appendices** | | |
| **S1 Appendix** | **Data extraction forms** | **59** |
| **S2 Appendix** | **Analysis of other systematic reviews on asymptomatic SARS-CoV-2** | **80** |

**S1 PRISMA Checklist. PRISMA Checklist 2020 and PRISMA Checklist for abstracts**

| Section and Topic | Item # | Checklist item | Location where item is reported |
|---|---|---|---|
| **TITLE** | | | |
| Title | 1 | Identify the report as a systematic review. | Page 1 |
| **ABSTRACT** | | | |
| Abstract | 2 | See the PRISMA 2020 for Abstracts checklist. | (See checklist below) |
| **INTRODUCTION** | | | |
| Rationale | 3 | Describe the rationale for the review in the context of existing knowledge. | Introduction page 6, paragraphs 1-2 |
| Objectives | 4 | Provide an explicit statement of the objective(s) or question(s) the review addresses. | Introduction page 7, paragraph 2 |
| **METHODS** | | | |
| Eligibility criteria | 5 | Specify the inclusion and exclusion criteria for the review and how studies were grouped for the syntheses. | Methods page 8, paragraph 3 |
| Information sources | 6 | Specify all databases, registers, websites, organisations, reference lists and other sources searched or consulted to identify studies. Specify the date when each source was last searched or consulted. | Methods page 8, paragraph 2 |
| Search strategy | 7 | Present the full search strategies for all databases, registers and websites, including any filters and limits used. | S1 Text |
| Selection process | 8 | Specify the methods used to decide whether a study met the inclusion criteria of the review, including how many reviewers screened each record and each report retrieved, whether they worked independently, and if applicable, details of automation tools used in the process. | Methods page 9, paragraph 4 |
| Data collection process | 9 | Specify the methods used to collect data from reports, including how many reviewers collected data from each report, whether they worked independently, any processes for obtaining or confirming data from study investigators, and if applicable, details of automation tools used in the process. | Methods page 9, paragraph 4. S1 Appendix. |
| Data items | 10a | List and define all outcomes for which data were sought. Specify whether all results that were compatible with each outcome domain in each study were sought (e.g. for all measures, time points, analyses), and if not, the methods used to decide which results to collect. | Methods page 9, paragraph 4. S1 Appendix |
| | 10b | List and define all other variables for which data were sought (e.g. participant and intervention characteristics, funding sources). Describe any assumptions made about any missing or unclear information. | Methods page 8, paragraph 3<br>Methods page 9, paragraph 4 |
| Study risk of bias assessment | 11 | Specify the methods used to assess risk of bias in the included studies, including details of the tool(s) used, how many reviewers assessed each study and whether they worked independently, and if applicable, details of automation tools used in the process. | Methods page 9, paragraph 5 |
| Effect measures | 12 | Specify for each outcome the effect measure(s) (e.g. risk ratio, mean difference) used in the synthesis or presentation of results. | Methods page 10, paragraph 6 |
| Synthesis methods | 13a | Describe the processes used to decide which studies were eligible for each synthesis (e.g. tabulating the study intervention characteristics and comparing against the planned groups for each synthesis (item #5)). | Methods page 10, paragraph 6 |
| | 13b | Describe any methods required to prepare the data for presentation or synthesis, such as handling of missing summary statistics, or data conversions. | Methods page 10, paragraph 6 |
| | 13c | Describe any methods used to tabulate or visually display results of individual studies and syntheses. | Methods page 10, paragraph 6 |
| | 13d | Describe any methods used to synthesize results and provide a rationale for the choice(s). If meta-analysis was performed, describe the model(s), method(s) to identify the presence and extent of statistical heterogeneity, and software package(s) used. | Methods page 10, paragraph 6 |

| | | | |
|---|---|---|---|
| | 13e | Describe any methods used to explore possible causes of heterogeneity among study results (e.g. subgroup analysis, meta-regression). | Methods page 10, paragraph 6 |
| | 13f | Describe any sensitivity analyses conducted to assess robustness of the synthesized results. | Methods page 10, paragraph 6 |
| Reporting bias assessment | 14 | Describe any methods used to assess risk of bias due to missing results in a synthesis (arising from reporting biases). | Methods page 10, paragraph 6 |
| Certainty assessment | 15 | Describe any methods used to assess certainty (or confidence) in the body of evidence for an outcome. | Does not apply |
| **RESULTS** | | | |
| Study selection | 16a | Describe the results of the search and selection process, from the number of records identified in the search to the number of studies included in the review, ideally using a flow diagram. | Results page 11, paragraph 1.<br>S1 Fig. |
| | 16b | Cite studies that might appear to meet the inclusion criteria, but which were excluded, and explain why they were excluded. | Results page 11, paragraph 1.<br>S1 Table |
| Study characteristics | 17 | Cite each included study and present its characteristics. | Results page 11, paragraph 1.<br>Table 1<br>S2 Table |
| Risk of bias in studies | 18 | Present assessments of risk of bias for each included study. | S3 Fig. |
| Results of individual studies | 19 | For all outcomes, present, for each study: (a) summary statistics for each group (where appropriate) and (b) an effect estimate and its precision (e.g. confidence/credible interval), ideally using structured tables or plots. | Results page 11-114, paragraph 2-4. Fig 1-2-3.<br>Table 1 |
| Results of syntheses | 20a | For each synthesis, briefly summarise the characteristics and risk of bias among contributing studies. | Results page 14, paragraph 5. |
| | 20b | Present results of all statistical syntheses conducted. If meta-analysis was done, present for each the summary estimate and its precision (e.g. confidence/credible interval) and measures of statistical heterogeneity. If comparing groups, describe the direction of the effect. | Results<br>Fig 1-2 |
| | 20c | Present results of all investigations of possible causes of heterogeneity among study results. | Results page 14, paragraph 6.<br>Table 2 |
| | 20d | Present results of all sensitivity analyses conducted to assess the robustness of the synthesized results. | Results page 14, paragraph 5.<br>S4 Table |
| Reporting biases | 21 | Present assessments of risk of bias due to missing results (arising from reporting biases) for each synthesis assessed. | Results page 14, paragraph 5.<br>S4 Table |
| Certainty of evidence | 22 | Present assessments of certainty (or confidence) in the body of evidence for each outcome assessed. | Does not apply |
| **DISCUSSION** | | | |
| Discussion | 23a | Provide a general interpretation of the results in the context of other evidence. | Discussion page 17, paragraph 1 |
| | 23b | Discuss any limitations of the evidence included in the review. | Discussion pages 17-18 paragraph 2 |
| | 23c | Discuss any limitations of the review processes used. | Discussion page 18, paragraph 2 |
| | 23d | Discuss implications of the results for practice, policy, and future research. | Discussion page 18-20, paragraphs 4-8 |

| OTHER INFORMATION | | | |
|---|---|---|---|
| Registration and protocol | 24a | Provide registration information for the review, including register name and registration number, or state that the review was not registered. | Methods page 7, paragraph 1 |
| | 24b | Indicate where the review protocol can be accessed, or state that a protocol was not prepared. | Methods page 7, paragraph 1 |
| | 24c | Describe and explain any amendments to information provided at registration or in the protocol. | Methods pages 7-8, paragraph 1 and 3 |
| Support | 25 | Describe sources of financial or non-financial support for the review, and the role of the funders or sponsors in the review. | This will appear in the article metadata |
| Competing interests | 26 | Declare any competing interests of review authors. | This will appear in the article metadata |
| Availability of data, code and other materials | 27 | Report which of the following are publicly available and where they can be found: template data collection forms; data extracted from included studies; data used for all analyses; analytic code; any other materials used in the review. | Methods page 10, paragraph 7. |

*From:* Page MJ, McKenzie JE, Bossuyt PM, Boutron I, Hoffmann TC, Mulrow CD, et al. The PRISMA 2020 statement: an updated guideline for reporting systematic reviews. BMJ 2021;372:n71. doi: 10.1136/bmj.n71

For more information, visit: http://www.prisma-statement.org/

PRISMA Checklist for abstracts

| **Section and Topic** | **Item #** | **Checklist item** | **Reported (Yes/No)** |
|---|---|---|---|
| **TITLE** | | | |
| Title | 1 | Identify the report as a systematic review. | Yes |
| **BACKGROUND** | | | |
| Objectives | 2 | Provide an explicit statement of the main objective(s) or question(s) the review addresses. | Yes |
| **METHODS** | | | |
| Eligibility criteria | 3 | Specify the inclusion and exclusion criteria for the review. | Yes |
| Information sources | 4 | Specify the information sources (e.g. databases, registers) used to identify studies and the date when each was last searched. | Yes |
| Risk of bias | 5 | Specify the methods used to assess risk of bias in the included studies. | Yes |
| Synthesis of results | 6 | Specify the methods used to present and synthesise results. | Yes |
| **RESULTS** | | | |
| Included studies | 7 | Give the total number of included studies and participants and summarise relevant characteristics of studies. | Yes |
| Synthesis of results | 8 | Present results for main outcomes, preferably indicating the number of included studies and participants for each. If meta-analysis was done, report the summary estimate and confidence/credible interval. If comparing groups, indicate the direction of the effect (i.e. which group is favoured). | Yes |
| **DISCUSSION** | | | |
| Limitations of evidence | 9 | Provide a brief summary of the limitations of the evidence included in the review (e.g. study risk of bias, inconsistency and imprecision). | Yes |
| Interpretation | 10 | Provide a general interpretation of the results and important implications. | Yes |
| **OTHER** | | | |
| Funding | 11 | Specify the primary source of funding for the review. | Yes |
| Registration | 12 | Provide the register name and registration number. | Yes |

#### S1 Text. Search strings

From: Living Evidence on COVID-19 (https://ispmbern.github.io/covid-19/living-review/collectingdata.html, accessed 06.07.2021)

We retrieve data from EMBASE via OVID, MEDLINE via PubMed, BioRxiv and MedRxiv.

**Search terms**

When searches are updated, references that are identified that were not in the database before, are inserted by date (**date_entrez**) they were indexed in remote database, the date they are inserted in OUR database is formatted as the '**strategydate**' (raw data is available here).

**01.05.2020**

EMBASE:

```
(SARS coronavirus/ or middle east respiratory syndrome/ or severe acute respiratory syndrome/ or (coronavirus* or corona virus* or HCoV* or ncov* or covid or covid19 or sars-cov* or sarscov* or Sars-coronavirus* or Severe Acute Respiratory Syndrome Coronavirus*).mp.) and 20191201:20301231.(dc).
```

**29.04.2020**

MEDLINE:

```
("coronavirus"[MH] OR "coronavirus infections"[MH] OR "coronavirus"[TW] OR "corona virus"[TW] OR "HCoV"[TW] OR "nCov"[TW] OR "covid"[TW] OR "covid19"[TW] OR "Severe Acute Respiratory Syndrome Coronavirus 2"[TW] OR "SARS-CoV2"[TW] OR "SARS-CoV 2"[TW] OR "SARS Coronavirus 2"[TW] OR "MERS-CoV"[TW]) AND (2019/1/1:3000[PDAT])
```

**01.04.2020**

From 01.04.2020, we retrieve the currate BioRxiv/MedRxiv dataset Link

**26.03.2020**

MEDLINE:

```
("Wuhan coronavirus" [Supplementary Concept] OR "COVID-19" OR SARS-CoV-2 OR "2019 ncov"[tiab] OR (("novel coronavirus"[tiab] OR "new coronavirus"[tiab]) AND (wuhan[tiab] OR 2019[tiab])) OR 2019-nCoV[All Fields] OR (wuhan[tiab] AND coronavirus[tiab]))
```

EMBASE:

```
(nCoV or 2019-nCoV or ((new or novel or wuhan) adj3 coronavirus) or covid19 or covid-19 or SARS-CoV-2).mp.
```

BioRxiv/MedRxiv:

```
ncov or corona or wuhan or COVID or SARS-CoV-2
```

With the kind support of the Public Health & Primary Care Library PHC, and following guidance of the Medical Library Association

**01.01.2020**

MEDLINE:

```
("Wuhan coronavirus" [Supplementary Concept] OR "COVID-19" OR "2019
ncov"[tiab] OR (("novel coronavirus"[tiab] OR "new coronavirus"[tiab]) AND
(wuhan[tiab] OR 2019[tiab])) OR 2019-nCoV[All Fields] OR (wuhan[tiab] AND
coronavirus[tiab])))))
```

EMBASE:

```
ncov OR (wuhan AND corona) OR COVID
```

BioRxiv/MedRxiv:

```
ncov or corona or wuhan or COVID
```

We retained publications that used the keywords listed below in the title or abstract.

"asymp*" OR "pre-symp*" OR "presymp*" OR "preclinical" OR "pre-clinical" OR "without symptoms" OR "no symptoms" OR "free of symptoms" OR "non-symp*" OR "nonsymp*" OR "symptom-free" OR "symptomfree"

#### S2 Text. Risk of Bias Tool

<table>
<tr><td colspan="3">RISK OF BIAS TOOL-ASYMPTOMATIC REVIEW</td></tr>
<tr><td colspan="3">Selection Bias</td></tr>
<tr><td colspan="3"><u>Reporting:</u> How was the target population described? (Extract it from the study)<br>Target population: The collection of individuals, items, measurements, etc., about which inferences are desired (Porta, 2014). For our review, our target population is comprised of all people or participants who are at risk of getting SARS-CoV-2, and after positive RT-PCR, they are followed up to assess if they develop symptoms or remain asymptomatic during infection.</td></tr>
<tr><td colspan="3">Question 1- Was the sample invited to participate a close or true representation of the target population?</td></tr>
<tr><td>High risk</td><td>Unclear</td><td>Low</td></tr>
<tr><td>High risk in case of the following:<br><br>• Volunteers: Authors only included people who volunteered to participate.<br><br>• Example: A large-scale study that requires voluntary participation instead of a random selection of participants.<br><br>• Sampling-based on symptoms.<br><br>• Other: Please explain further.</td><td>• No information about sampling strategy.</td><td>• All eligible participants were included from a random sample.<br><br>• Contact tracing studies that included all contacts from index cases positive for SARS-CoV-2.<br><br>• All potential people who could be positive for SARS-CoV-2 were included in a screening study.<br><br>Examples:<br><br>• A study where all people had to be tested after traveling. Those who were positive were quarantined in a medical facility.<br><br>• All people who visited a facility where an outbreak occurred.<br><br>• All Residents in a nursing facility</td></tr>
<tr><td colspan="3"><u>Reporting:</u> Was the response rate from the eligible population provided?<br>Yes<br>No<br>Sometimes the authors report the participation or response rate. However, be very careful about the numbers provided by the authors</td></tr>
</table>

| |
|---|
| Example<br>• Eligible population: All passengers who were screened before taking a flight in the United States.<br>• Eligible Participants: All people who got tested for SARS-CoV-2 independently of the result. |
| If not, can you calculate the response rate? You can try to find if the authors report the number of people who met the inclusion criteria and/ or were invited and how many were finally included in the study (positive and negative for SARS-CoV-2 ).<br><br>$\text{Response Rate} = \frac{Number\ of\ individuals\ who\ participated\ in\ the\ study}{Number\ of\ elibigle\ participants}\ X\ 100$<br><br>**Example**<br>**Example**<br>• *Eligible participants:* All passengers before taking a flight in the United States.<br>• *Individuals who participated in the study:* All passengers who got tested for SARS-CoV-2 independently of the result.<br><br>(Number of people who participated in the study/number of eligible participants)<br>Yes<br>No<br>Response Rate:<br><br>Num and denominator% |
| Question 2 - The characteristics of non-respondents, if any, who were eligible are similar to those who participated in the study? |

| | | |
|---|---|---|
| *Response rate bias occurs when people who do not take part in a study differ systematically from those who take part in ways that are associated with the condition of interest, even if those invited come from a random selection.*<br><br>We will not provide an exact cut-point to say there is a risk of response rate bias. We want to assess if there are differences among those who participated and those who did not participate in the study | | |
| **High** | **Unclear** | **Low** |
| • Respondents and non-respondents are different, and the participation might be related to being symptomatic or asymptomatic.<br><br>• Other: Please explain further. | • They might be similar, but no information was provided. | • The characteristics between respondents and non-respondents are similar.<br><br>• There were no non-respondents, the response rate is 100% |
| **Information Bias** | | |
| Reporting: How was symptomatic and asymptomatic defined? | | |
| Based on the reporting question<br>Question 3 - Was the assessment of symptoms status adequate? In this question, we would like to find the specific type of symptoms assessed in each study (for the type of symptoms commonly present in patients with COVID-19, see link). Please be aware of the appendixes in the supplementary material. In many cases, the list of symptoms is available in this section. | | |
| **High** | **Unclear** | **Low** |
| • The list of assessed symptoms is limited; for example, the study only presents information about flu-like symptoms.<br><br>• Other: Please explain further. | • There is no clear information about symptoms; the authors mention only symptomatic and asymptomatic without further description. | The authors assessed the symptom status based on a well-defined list of symptoms.<br><br>See link WHO-list of symptoms |
| https://www.who.int/emergencies/diseases/novel-coronavirus-2019/question-and-answers-hub/q-a-detail/coronavirus-disease-covid-19 | | |

| Question 4- Based on the method symptoms were collected, is there a risk of recall bias?<br>Recall bias: Occurs when the condition has been measured through surveys or questionnaires that rely on memory to provide information on the condition of interest.<br><br>We are aware that symptoms are a subjective report, but we rely on the participant's memory to answer our research question. However, we want to assess if researchers asked participants or located the information about symptoms with enough frequency to minimize recall bias. Please see the options for high, low, or unclear. | | |
|---|---|---|
| **High** | **Unclear** | **Low** |
| Symptom status was assessed with a delay of one or two weeks.<br><br>• Example: Participants were asked, 'Have you had any symptoms or the following symptoms during the past two weeks?' | • There is no clear information about the method for collecting symptoms. | • Symptom status was collected frequently during the follow-up. (i.e., daily, two times per week)<br><br>• Symptom status was assessed retrospectively in clinical charts, and the researchers stated that symptom status was frequently evaluated (i.e., daily, two times per week). |
| **INFORMATION BIAS - Misclassification** | | |
| Reporting: In the study, how did the authors report the follow-up of symptom status? You can select or more options<br>✓ 14 days after exposure<br>✓ Until one or more negative PCR<br>✓ Seven days after testing and 14 days after exposure<br>✓ More than one of the mentioned above<br>✓ Longer follow-up | | |
| Was the viral load reported?<br>Yes<br>No<br><br>If yes, please provide the information about it CT. Add definition<br>*CT: Cycle threshold | | |

| Question 5- Is there a risk that the asymptomatic status was misclassified because of the follow-up? This question is not about the type of symptoms. It is about the follow-up period of symptoms among those who were positive for SARS-CoV-2 taking into account the course of the infection. | | |
|---|---|---|
| **High** | **Unclear** | **Low** |
| • Follow-up 7 days after testing<br>• Other: Please explain further. | • Follow-up 14 days after the last possible exposure | • Until negative PCR<br>• 14 after exposure and 7 days after testing<br>• A Longer follow-up |
| **Selective or incomplete reporting of outcome (asymptomatic status)** | | |
| Reporting<br>The authors provide information about the symptom status of all positive cases<br>Yes<br>No | | |
| Question 6- Based on your previous answer<br>Is there a risk of incomplete or selective reporting of symptoms status among those positive for SARS-CoV-2? | | |
| **High** | **Unclear** | **Low** |
| • There is missing information about the final symptom status of a proportion of participants. | • The numbers are not clearly reported | • All participants were followed up for symptom status and were included in the analysis. |
| Based on the risk of bias assessment, please provide some thoughts about the overall assessment of the study. Is there a risk of over or under-estimating the proportion of the truly asymptomatic population?<br><br>Please select one of the following and write down your comments about it | | |

**S1 Table. Studies included in version 3.0 and excluded in version 4.0 of the living systematic review**

| Author | Reason for Exclusion |
|---|---|
| **Contact investigations** | |
| Tong, ZD | Case series of already diagnosed cases |
| Huang, R | Contact investigation of a single family or individual |
| Jiang, XL | Contact investigation of a single family or individual |
| Jiang, X | Contact investigation of a single family or individual |
| Liao, J | The study only included diagnosed cases |
| Hu, Z | The study only included diagnosed cases |
| Luo, SH | Contact investigation of a single family or individual |
| Chan, JF | Contact investigation of a single family or individual |
| Ye, F | Contact investigation of a single family or individual |
| Bai, Y | Contact investigation of a single family or individual |
| Luo, Y | Contact investigation of a single family or individual |
| Zhang, J | Contact investigation of a single family or individual |
| Zhang, B | Contact investigation of a single family or individual |
| Huang, L | Contact investigation of a single family or individual |
| Qian, G | Contact investigation of a single family or individual |
| Gao, Y | Case series that do not enroll consecutive patients |
| **Contact investigations, aggregated** | |
| Wang, Z | Not all contacts were tested for SARS-CoV-2 |
| Yang, R | The study only included diagnosed cases |
| Bi, Q | Asymptomatic status only ascertained at start |
| **Outbreak investigations** | |
| Roxby, AC | Inadequate follow-up, unclear information about the last possible exposure. |
| Solbach, W | The study only included diagnosed cases |

| | |
|---|---|
| Mizumoto, K | Data included in another publication |
| Tian, S | Preprint of published article (6685) |
| Pham, TQ | Preprint of published article (1904) |
| **Screening** | |
| Arima, Y | Same population reported twice |
| Lytras, T | Only included pre-symptomatic patients |
| Lombardi, A | Preprint of published article (1264) |
| **Hospitalised adults** | |
| Pongpirul, WA | The study only included diagnosed cases |
| Qiu, C | The study only included diagnosed cases |
| Zou L | The study only included diagnosed cases |
| Zhou, R | The study only included diagnosed cases |
| Chang, MC | The study only included diagnosed cases |
| Zhou, X | The study only included diagnosed cases |
| Angelo Vaira, L | The study only included diagnosed cases |
| Wang, X | The study only included diagnosed cases |
| Xu, T | The study only included diagnosed cases |
| Tabata, S | The study only included diagnosed cases |
| Noh, JY | The study only included diagnosed cases |
| Kumar, R | The study only included diagnosed cases |
| Meng, H | Only included pre-symptomatic patients |
| Zhang, Z | Only included pre-symptomatic patients |
| Al-Shamsi, HO | Only included pre-symptomatic patients |
| Wang, Y1 | Only included pre-symptomatic patients |
| **Hospitalised children** | |
| See, KC | The study only included diagnosed cases |
| Tan, YP | The study only included diagnosed cases |
| Tan, X | The study only included diagnosed cases |
| Melgosa, M | The study only included diagnosed cases |

| | |
|---|---|
| Wu, HP | Inadequate follow-up. |
| Song, W | The study only included diagnosed cases |
| Bai, K | The study only included diagnosed cases |
| Xu, H | The study only included diagnosed cases |
| Qiu, H | The study only included diagnosed cases |
| Lu, Y | The study only included diagnosed cases |
| **Hospitalised adults and children** | |
| Merza, MA | The study only included diagnosed cases |
| Yongchen, Z | The study only included diagnosed cases |
| Ma, Y | The study only included diagnosed cases |
| Kim, SE | The study only included diagnosed cases |
| Choe, PG | The study only included diagnosed cases |
| Sharma, AK | The study only included diagnosed cases |
| Zhang, W3 | The study only included diagnosed cases |
| Alshami, AA | The study only included diagnosed cases |
| Kong, W | The study only included diagnosed cases |
| Wang, Y2 | The study only included diagnosed cases |
| **Mathematical Models** | |
| Ganyani, T | Data included in another publication |
| Kim, Y | Preprint of published article (2034, first author Chun) |
| Emery | Preprint of published article (2325) |
| Casey | Mathematical model not in review scope - published version did not include pooled estimate of presymptomatic transmission |

**S2 Table. Characteristics of studies reporting on proportion of asymptomatic SARS-CoV-2 infections (review question 1 and review question 2)**

| Study [a] | Location[b] | Total SARS-CoV-2, n | People with asymptomatic SARS-CoV-2 infection | | | Follow-up method[d] |
|---|---|---|---|---|---|---|
| | | | n | Sex | Age[c] | |
| **Contact and outbreak investigations** | | | | | | |
| Pirnay JP, 2020 [1] | Belgium | 4 | 2 | 0 F, 2 M | 28.5<br>IQR 25-37 | 1 |
| Cardillo L, 2021 [Healthcare workers] [2] | Italy, Campania | 4 | 2 | 11 F, 9 M | 78<br>IQR 72-85 | 4 |
| Garibaldi PMM, 2021 [Staff] [3] | Brazil, Sao Paulo State | 8 | 0 | NR | NR | 1, 2, 4 |
| Corcorran MA, 2020 [4] | United States of America, Washington | 8 | 3 | NR | NR | 2, 3 |
| Yang N, 2020 [5] | China, Xiaoshan | 10 | 2 | 1 F, 1 M | 26 | 1, 2 |
| Hijnen D, 2020 [6] | Germany, Munich | 11 | 1 | 0 F, 1 M | 49 | 1, 2 |
| Schwierzeck V, 2020 [7] | Germany, Muenster | 11 | 2 | NR | NR | 2 |
| Danis K, 2020 [8] | France | 12 | 1 | NR | NR | 1, 2, 4 |
| Garibaldi PMM, 2021 [3] | Brazil, Sao Paulo State | 12 | 1 | NR | NR | 1, 2, 4 |
| Zhang W, 2020 [9] | China, Guangzhou | 12 | 4 | NR | NR | 1, 2, 3 |
| Romao VC, 2020 [10] | Portugal, Lisbon | 14 | 0 | NR | NR | 3 |
| Orsi A, 2021 [11] | Italy, Genoa | 14 | 13 | 9 F, 4 M | 91<br>IQR 84-93 | 1, 4 |
| Böhmer MM, 2020 [12] | Germany, Bavaria | 16 | 1 | NR | NR | 1, 2 |

| | | | | | | |
|---|---|---|---|---|---|---|
| Dora AV, 2020 [13] | United States of America, Los Angeles | 16 | 6 | 0 F, 6 M | 75<br>IQR 72-75 | 3 |
| Yau K, 2020 [14] | Canada, Toronto | 20 | 7 | NR | NR | 2, 4 |
| Cheng HY, 2020 [15] | Other, Taiwan | 22 | 4 | NR | NR | 1 |
| Redditt V, 2020 [16] | Canada, Toronto | 24 | 3 | NR | NR | 2, 4 |
| Tian S, 2021 [17] | China, Shandong | 24 | 7 | NR | NR | 2, 3 |
| Harada S, 2020 [Patients] [18] | Japan, Tokyo | 24 | 8 | NR | NR | 2, 4 |
| Park JH, 2020 [19] | South Korea | 28 | 4 | NR | NR | 1, 2 |
| Patel MC, 2020 [20] | United States of America, Illinois | 35 | 13 | NR | NR | 2, 4 |
| Brandstetter S, 2020 [21] | Germany, Regensburg | 36 | 2 | NR | NR | 2, 4 |
| Kittang BR, 2020 [22] | Norway, Bergen | 40 | 0 | NR | NR | 1, 2 |
| Pavli A, 2020 [23] | Greece | 46 | 7 | NR | NR | 1, 2 |
| Yousaf AR, 2020 [24] | United States of America, Utah | 47 | 0 | NR | NR | 2, 4 |
| Arons MM, 2020 [25] | United States of America, Seattle, WA | 47 | 3 | NR | NR | 2 |
| Wu J, 2020 [26] | China, Zhuhai | 48 | 5 | NR | NR | 1, 2, 4 |
| Harada S, 2020 [Healthcare workers] [18] | Japan, Tokyo | 49 | 25 | NR | NR | 2, 4 |
| Xie W, 2021 [27] | China, Beijing | 53 | 4 | NR | NR | 2 |
| Ladhani SN, 2020 | United Kingdom, London | 53 | 26 | NR | NR | 2, 4 |

| | | | | | | |
|---|---|---|---|---|---|---|
| [Healthcare workers] [28] | | | | | | |
| van den Besselaar JH, 2021 [Healthcare workers] [29] | Netherlands, South Holland | 54 | 1 | NR | NR | 2, 4 |
| Schmitt J, 2021 [30] | Côte d'Ivoire | 54 | 18 | NR | NR | 1, 2, 4 |
| Gettings JR, 2021 [31] | United States of America, Georgia | 55 | 31 | NR | NR | 1, 2, 4 |
| Plucinski MM, 2020 [32] | Japan | 66 | 14 | NR | NR | 2 |
| Njuguna H, 2020 [33] | United States of America, Louisiana | 71 | 29 | NR | NR | 1, 2 |
| Jones A, 2021[34] | United States of America, Vermont | 87 | 24 | NR | NR | 1, 2 |
| Cardillo L, 2021 [Patients] [2] | Italy, Campania | 91 | 20 | 11 F, 9 M | 78<br>IQR 72-85 | 4 |
| Park SY, 2020 [35] | South Korea, Seoul | 95 | 4 | NR | NR | 2, 4 |
| Taylor J, 2020 [Healthcare personnel] [36] | United States of America, Minnesota | 99 | 9 | NR | NR | 2, 4 |
| Grijalva CG, 2020 [37] | United States of America, Tennessee and Wisconsin | 102 | 34 | NR | NR | 2 |
| Ladhani SN, 2020 [Residents] [28] | United Kingdom, London | 105 | 46 | NR | NR | 2, 4 |
| Paleker M, 2021 [38] | South Africa | 112 | 41 | NR | NR | 4 |
| van den Besselaar JH, 2021 [Residents] [29] | Netherlands, South Holland | 113 | 7 | NR | NR | 2, 4 |

| | | | | | | |
|---|---|---|---|---|---|---|
| Graham N, 2020 [39] | United Kingdom, London | 126 | 46 | NR | NR | 2 |
| Taylor J, 2020 [Residents] [36] | United States of America, Minnesota | 127 | 7 | NR | NR | 2, 4 |
| Luo L2, 2020 [40] | China, Guangzhou | 127 | 8 | NR | NR | 1, 4 |
| Shi Q, 2020 [41] | China, Wanzhou District | 183 | 60 | NR | NR | 3 |
| Pham QT, 2020 [42] | Vietnam | 208 | 89 | NR | 31<br>IQR 23-45 | 2 |
| Hurst JH, 2020 [43] | United States of America, North Carolina | 293 | 87 | NR | NR | 2 |
| Kennelly SP, 2020 [Nursing home staff] [44] | Ireland | 395 | 97 | NR | NR | 2 |
| Lee JY, 2020 [45] | South Korea, Daegu | 694 | 80 | NR | NR | 2 |
| Kennelly SP, 2020 [Nursing home residents] [44] | Ireland | 710 | 193 | NR | NR | 2 |
| Kasper MR, 2020 [46] | United States of America | 1,271 | 572 | NR | NR | 2, 4 |
| **Screening in community, institutional and occupational settings** | | | | | | |
| Hoehl S, 2020 [47] | Germany, Germersheim | 2 | 1 | 0 F, 1 M | 58 | 2 |
| Tanacan A, 2020 [48] | Turkey, Ankara | 3 | 0 | NR | NR | 3 |
| Jeffery-Smith A, 2021 [Staff] [49] | United Kingdom, London | 3 | 2 | NR | NR | 3 |
| Fisher MJ, 2021 [50] | United States of America, New York | 4 | 3 | 0 F, 3 M | 78<br>IQR 77.5 | 2 |

| | | | | | | |
|---|---|---|---|---|---|---|
| Chang L, 2020 [51] | China, Wuhan | 4 | 2 | 0 F, 2 M | 45<br>IQR 37-53 | 2, 3 |
| Berghoff AS, 2020 [52] | Austria, Vienna | 4 | 2 | NR | 53<br>IQR 43-63 | 1, 2, 3 |
| Vohra LM, 2021 [Undergoing chemotherapy] [53] | Pakistan | 4 | 2 | NR | NR | 2, 3, 4 |
| Pamplona J, 2021 [54] | Spain, Girona | 5 | 2 | 2 F, 0 M | 72<br>IQR 63-81 | 2, 4 |
| Rauch JN, 2021 [55] | United States of America, California | 6 | 2 | NR | NR | 2, 4 |
| AbdulRahman A, 2020 [56] | Bahrain | 6 | 3 | 0 F, 3 M | 25 | 2, 3 |
| Viñuela MC, 2020 [57] | Spain, Madrid | 8 | 8 | 8 F, 0 M | 32 | 3 |
| Bender WR, 2020 [58] | United States of America, Philadelphia, PA | 8 | 6 | 6 F, 0 M | NR | 2, 4 |
| Lalani T, 2021 [59] | United States of America, New York City | 8 | 4 | NR | NR | 1, 2, 4 |
| van Buul LW, 2020 [Healthcare workers] [60] | Netherlands | 9 | 0 | NR | NR | 2, 4 |
| Ferreira VH, 2021 [61] | Canada, Toronto | 9 | 5 | 5 F, 0 M | 33<br>IQR 4.75 | 2, 4 |
| Vohra LM, 2021 [Presurgical patients] [53] | Pakistan | 10 | 10 | NR | NR | 2, 3, 4 |
| Theuring S, 2021 [School students and staff] [62] | Germany, Berlin | 10 | 2 | NR | NR | 2, 4 |
| Varnell C, 2021 [63] | United States of America | 10 | 5 | NR | NR | 2 |

| | | | | | | |
|---|---|---|---|---|---|---|
| Morgan SC, 2021 [64] | United States of America, San Diego | 11 | 2 | NR | NR | 2, 4 |
| Kutsuna S, 2020 [65] | Japan, Tokyo | 11 | 3 | 1 F, 2 M | NR | 2, 3 |
| Kirshblum SC, 2020 [66] | United States of America, New Jersey | 12 | 2 | NR | NR | 2, 4 |
| Haidar G, 2021 [67] | United States of America, Pennsylvania | 11 | 9 | 6 F, 3 M | 54<br>IQR 74-31 | 2, 4 |
| Jeffery-Smith A, 2021 [Residents] [49] | United Kingdom, London | 13 | 6 | NR | NR | 3 |
| Theuring S, 2021 [Household members] [62] | Germany, Berlin | 14 | 0 | NR | NR | 2, 4 |
| Hwang, 2021 [68] | United States of America | 14 | 8 | NR | NR | 2, 4 |
| Isoldi S, 2021 [69] | Italy, Rome | 15 | 4 | NR | NR | 3, 4 |
| van Buul LW, 2020 [Nursing home residents] [60] | Netherlands | 16 | 3 | NR | NR | 2, 4 |
| Han X, 2020 [70] | China, Wuhan | 17 | 17 | 8 F, 9 M | 30<br>IQR 27-30 | 2 |
| Wadhwa A, 2020 [71] | United States of America, Chicago | 17 | 6 | NR | NR | 2 |
| Balestrini S, 2020 [72] | United Kingdom, London | 17 | 11 | NR | NR | 3 |
| Alshahrani MS, 2020 [73] | Saudi Arabia, Alkhobar | 18 | 12 | NR | NR | 3 |
| Maki G, 2020 [74] | United States of America, Detroit, Michigan | 18 | 16 | 9 F, 7 M | mean 50.7 | 2, 4 |

| | | | | | | |
|---|---|---|---|---|---|---|
| Stock AD, 2020 [75] | United States of America, New York City | 19 | 6 | NR | NR | 1, 2 |
| Bogani G, 2020 [76] | Italy, Lombardy | 19 | 10 | 10 F, 0 M | NR | 3 |
| Edelstein M, 2020 [77] | United Kingdom, London | 20 | 4 | NR | NR | 3, 4 |
| Fakhim H, 2021 [78] | Iran, Isfahan | 21 | 14 | NR | NR | 2, 3 |
| Green R, 2021 [79] | United Kingdom, Liverpool | 22 | 22 | 15 F, 7 M | 80<br>IQR 19-106 | 2, 4 |
| Martins Machado C, 2020 [80] | Brazil | 22 | 5 | NR | NR | 2, 4 |
| Khondaker T, 2021 [81] | Bangladesh, Dhaka | 26 | 7 | NR | NR | 2, 4 |
| Laws RL, 2021 [82] | Uganda, Kampala | 28 | 25 | NR | NR | 2, 4 |
| Rivett L, 2020 [83] | United Kingdom, Cambridge | 30 | 5 | NR | NR | 2 |
| Starling A, 2020 [84] | United Kingdom, Essex | 31 | 29 | NR | NR | 2, 4 |
| Rincon A, 2020 [85] | Spain, Barcelona | 35 | 9 | 2 F, 7 M | 76.67<br>SD 13.86 | 1, 2, 4 |
| Malagón-Rojas J, 2020 [86] | Colombia, Bogota | 35 | 11 | NR | NR | 2, 4 |
| Pizarro-Sánchez MS, 2021 [87] | Spain, Madrid | 38 | 5 | 2 F, 3 M | NR | 2, 4 |
| Hogan CA, 2021 [88] | United States of America, California | 38 | 20 | 13 F, 7 M | 41<br>IQR 32.5-47 | 2, 4 |
| Treibel TA, 2020 [89] | United Kingdom, London | 44 | 12 | NR | NR | 2, 4 |
| Tan-Loh J, 2021 [90] | Malaysia, Teluk Intan | 46 | 6 | NR | NR | 2, 3, 4 |
| Letizia AG, 2020 [91] | United States of America, South Carolina | 51 | 46 | NR | NR | 1, 2 |

| | | | | | | |
|---|---|---|---|---|---|---|
| Migisha R, 2020 [92] | Uganda | 54 | 20 | NR | NR | 2, 4 |
| Patel MR, 2021 [93] | India | 55 | 43 | NR | NR | 4 |
| Aslam A, 2020 [94] | United States of America, New York | 65 | 38 | NR | NR | 2, 4 |
| Marossy A, 2020 [95] | United Kingdom, London | 67 | 46 | NR | NR | 2, 3 |
| London V, 2020 [96] | United States of America, New York City | 68 | 22 | 22 F, 0 M | 30.5<br>IQR 24.5-34.8 | 2, 4 |
| Lavezzo E, 2020 [97] | Italy, Veneto | 73 | 29 | NR | NR | 2 |
| Chamie G, 2020 [98] | United States of America, San Francisco | 81 | 23 | NR | NR | 2, 4 |
| Meyers KJ, 2021 [99] | United States of America, Indianapolis | 86 | 67 | NR | NR | 2, 4 |
| Smith E, 2020 [100] | United Kingdom, Norfolk | 103 | 42 | NR | NR | 2, 4 |
| Weinbergerova B, 2021 [101] | Czech Republic, Brno | 105 | 6 | NR | NR | 3 |
| Wi YM, 2020 [102] | South Korea, Gyeongsangnam-do province | 111 | 7 | NR | NR | 3 |
| Esteban I, 2021 [103] | Argentina, Buenos Aires | 113 | 75 | NR | NR | 4 |
| Nunes MC, 2021 [104] | South Africa, Soweto | 115 | 14 | NR | NR | 3 |
| Turunen T, 2021 [105] | Finland | 127 | 23 | 0 F, 23 M | NR | 1 |
| Hcini N, 2020 [106] | French Guyana, West French Guiana territory | 137 | 87 | 87 F, 0 M | NR | 2, 4 |
| Wong J, 2020 [107] | Brunei | 138 | 16 | NR | NR | 2, 3 |
| Lombardi A, 2020 [108] | Italy, Lombardy | 139 | 17 | NR | NR | 1, 2, 3 |

| | | | | | | |
|---|---|---|---|---|---|---|
| Shi SM, 2020 [109] | United States of America, Boston, MA | 146 | 21 | NR | NR | 2, 4 |
| Andrikopoulou M, 2020 [110] | United States of America, New York | 158 | 46 | 46 F, 0 M | NR | 2 |
| Blain H, 2021 [111] | France | 161 | 14 | NR | NR | 1, 2, 4 |
| Say D, 2021 [112] | Australia, Melbourne | 171 | 61 | NR | NR | 4 |
| Beiting KJ, 2021 [113] | United States of America, Chicago, IL | 172 | 50 | NR | NR | 2, 4 |
| Uçkay, I, 2021 [114] | Switzerland, Zurich | 175 | 71 | NR | NR | 2, 4 |
| Eythorsson E, 2020 [115] | Iceland | 178 | 25 | NR | NR | 2, 4 |
| Cariani L, 2020 [116] | Italy, Milan | 182 | 32 | NR | NR | 2, 3 |
| Al-Qahtani M, 2020 [117] | Bahrain | 188 | 116 | NR | NR | 2, 3, 4 |
| Adhikari EH, 2020 [118] | United States of America, Texas | 252 | 98 | 98 F, 0 M | NR | 2, 4 |
| Hussain A, 2020 [119] | Pakistan, Karachi | 266 | 54 | NR | NR | 3 |
| Cao S, 2020 [120] | China, Wuhan | 300 | 300 | 168 F, 132 M | NR | 1, 3 |
| Marcus JE, 2021 [121] | United States of America, Texas | 403 | 199 | NR | NR | 2, 4 |
| Ghinai I, 2020 [122] | United States of America, Chicago | 406 | 293 | NR | NR | 2, 4 |
| Mahajan NN, 2020 [123] | India, Mumbai | 467 | 58 | NR | NR | 3 |
| Uysal E, 2021[124] | Turkey | 684 | 64 | 42 F, 22 M | 59.4 SD 12 | 2, 4 |

| | | | | | | |
|---|---|---|---|---|---|---|
| Almazeedi S, 2020 [125] | Kuwait | 1,096 | 473 | NR | NR | 3 |
| Hall VJ, 2021 [126] | United Kingdom | 1,704 | 293 | NR | NR | 2, 4 |
| Malhotra S, 2021 [127] | India | 1,729 | 1,272 | NR | NR | 2 |
| Abraha HE, 2021 [128] | Ethiopia, Tigray | 2,617 | 1,935 | NR | NR | 2, 4 |
| Ren R, 2021 [129] | China | 3,103 | 1,612 | 378 F, 1234 M | NR | 2, 4 |
| White EM, 2020 [130] | United States of America | 5,403 | 2,194 | NR | NR | 2, 4 |
| Bender JK, 2021[e] [131] | Germany | 98 | 26 | NR | NR | 1,3 |
| Wu P, 2021[e] [132] | China | 4214 | 12 | 10 F, 2 M | NR | 2 |

SARS-CoV-2, severe acute respiratory syndrome coronavirus 2; NR, not reported; F, female; M, male; IQR, interquartile range.

[a] See reference list of studies included for question 1 and question 2. Reference numbers differ from the main text.

[b]Location is reported as described in the study.

[c] Median and interquartile range (IQR) or mean and standard deviation (SD)

[d] Follow-up recorded according to study protocol (1: 14 days after last possible exposure; 2: 7 days after diagnosis; 3:>7 days after diagnosis; 4: until negative RT-PCR result).

[e]Studies only included for question 2.

15. Cheng HY, Jian SW, Liu DP, Ng TC, Huang WT, Lin HH, et al. Contact Tracing Assessment of Covid-19 Transmission Dynamics in Taiwan and Risk at Different Exposure Periods before and after Symptom

Onset. JAMA Intern Med. 2020;180(9):1156-63. Epub 2020/05/02. https://doi.org/10.1001/jamainternmed.2020.2020. PubMed PMID: 32356867; PubMed Central PMCID: 7195694.

Epub 20210212. https://doi.org/10.1002/jmv.26847. PubMed PMID: 33538342; PubMed Central PMCID: PMC8014049.

2020/12/07. https://doi.org/10.1016/j.yebeh.2020.107602. PubMed PMID: 33279440; PubMed Central PMCID: 7643621.

96. London V, McLaren R, Jr., Atallah F, Cepeda C, McCalla S, Fisher N, et al. The Relationship between Status at Presentation and Outcomes among Pregnant Women with Covid-19. Am J Perinatol.

2020;37(10):991-4. Epub 2020/05/20. https://doi.org/10.1055/s-0040-1712164. PubMed PMID: 32428964; PubMed Central PMCID: 7416203.

https://doi.org/10.1001/jamainternmed.2020.5664. PubMed PMID: 33074318; PubMed Central PMCID: 7573793.

131. Bender JK, Brandl M, Höhle M, Buchholz U, Zeitlmann N. Analysis of Asymptomatic and Presymptomatic Transmission in Sars-Cov-2 Outbreak, Germany, 2020. Emerging infectious diseases. 2021;27(4):1159-63. Epub 2021/02/18. https://dx.doi.org/10.3201%2Feid2704.204576. PubMed PMID: 33600301.

132. Wu P, Liu F, Chang Z, Lin Y, Ren M, Zheng C, et al. Assessing Asymptomatic, Presymptomatic, and Symptomatic Transmission Risk of Severe Acute Respiratory Syndrome Coronavirus 2. Clin Infect Dis. 2021;73(6):e1314-e20. https://doi.org/10.1093/cid/ciab271. PubMed PMID: 33772573; PubMed Central PMCID: PMC8083716.

**S3 Table. Location of studies contributing data to review question 1**

| Country | Total SARS-CoV-2, n | Total asymptomatic SARS-CoV-2, n | Total number of studies |
|---|---|---|---|
| United States of America | 9,725 | 4,004 | 37 |
| United Kingdom | 2,338 | 590 | 12 |
| China | 3,881 | 2,021 | 11 |
| Italy | 537 | 127 | 7 |
| Germany | 100 | 9 | 6 |
| South Korea | 928 | 95 | 4 |
| Spain | 86 | 24 | 4 |
| Canada | 53 | 15 | 3 |
| India | 2,251 | 1,373 | 3 |
| Japan | 150 | 50 | 3 |
| Bahrain | 194 | 119 | 2 |
| Brazil | 42 | 6 | 2 |
| France | 173 | 15 | 2 |
| Netherlands | 192 | 11 | 2 |
| Pakistan | 280 | 66 | 2 |
| South Africa | 227 | 55 | 2 |
| Turkey | 687 | 64 | 2 |
| Uganda | 82 | 45 | 2 |
| Argentina | 113 | 75 | 1 |
| Australia | 171 | 61 | 1 |
| Austria | 4 | 2 | 1 |
| Bangladesh | 26 | 7 | 1 |
| Belgium | 4 | 2 | 1 |
| Brunei | 138 | 16 | 1 |

| | | | |
|---|---|---|---|
| Colombia | 35 | 11 | 1 |
| Czech Republic | 105 | 6 | 1 |
| Ethiopia | 2,617 | 1,935 | 1 |
| Finland | 127 | 23 | 1 |
| French Guyana | 137 | 87 | 1 |
| Greece | 46 | 7 | 1 |
| Iceland | 178 | 25 | 1 |
| Iran | 21 | 14 | 1 |
| Ireland | 1,105 | 290 | 1 |
| Ivory Coast | 54 | 18 | 1 |
| Kuwait | 1,096 | 473 | 1 |
| Malaysia | 46 | 6 | 1 |
| Norway | 40 | 0 | 1 |
| Other | 22 | 4 | 1 |
| Portugal | 14 | 0 | 1 |
| Saudi Arabia | 18 | 12 | 1 |
| Switzerland | 175 | 71 | 1 |
| Vietnam | 208 | 89 | 1 |

**S4 Table. Summary of findings from subgroup analyses in studies estimating the proportion of asymptomatic SARS-CoV-2 infections**

| | Contact and outbreak investigations | | | | | | Screening of defined population | | | | | |
|---|---|---|---|---|---|---|---|---|---|---|---|---|
| **Domain** | **n[a]** | **Summary (95% CI)** | **Prediction interval** | **$I^2$, %** | **$\tau^2$** | **Subgroup difference, p value** | **n[b]** | **Summary (95% CI)** | **Prediction interval** | **$I^2$, %** | **$\tau^2$** | **Subgroup difference p value** |
| **Selection bias[b]** | | | | | | | | | | | | |
| Low risk | 27 | 0.24 (0.17-0.33) | 0.04-0.74 | 84 | 1.05 | 0.097 | 27 | 0.46 (0.33-0.59) | 0.05-0.94 | 98 | 1.85 | 0.597 |
| Unclear/ high risk | 26 | 0.15 (0.10-0.23) | 0.02-0.68 | 88 | 1.36 | | 61 | 0.41 (0.31;0.52) | 0.03-0.94 | 97 | 2.45 | |
| **Information bias, assessment of symptoms defining status[a]** | | | | | | | | | | | | |
| Low risk | 12 | 0.18 (0.08-0.35) | 0.01-0.86 | 85 | 2.07 | 0.794 | 21 | 0.29 (0.20-0.42) | 0.03-0.83 | 98 | 1.33 | 0.026 |
| Unclear/ high risk | 41 | 0.20 (0.15;0.26) | 0.03-0.67 | 86 | 1.07 | | 67 | 0.47 (0.37;0.57) | 0.04-0.95 | 97 | 2.47 | |
| **Information bias, misclassification based on follow-up[a]** | | | | | | | | | | | | |
| Low risk | 39 | 0.18 (0.13-0.26) | 0.02-0.76 | 88 | 1.64 | 0.241 | 68 | 0.41 (0.32-0.51) | 0.03-0.94 | 97 | 2.31 | 0.554 |
| Unclear/ high risk | 14 | 0.24 (0.18-0.32) | 0.07-0.55 | 88 | 0.35 | | 20 | 0.47 (0.31-0.63) | 0.04-0.95 | 97 | 1.95 | |
| **Selective reporting bias[a]** | | | | | | | | | | | | |
| Low risk | 47 | 0.20 (0.15-0.26) | 0.03-0.69 | 90 | 1.18 | 0.758 | 79 | 0.44 (0.35-0.53) | 0.03-0.95 | 97 | 2.40 6 | 0.242 |
| Unclear/ high risk | 6 | 0.17 (0.06-0.38) | 0.00-0.92 | 92 | 1.74 | | 9 | 0.32 (0.18-0.50) | 0.03-0.87 | 97 | 1.08 | |
| **All domains** | | | | | | | | | | | | |
| Low risk | 6 | 0.20 (0.09-0.39) | 0.01-0.85 | 92 | 1.01 | 0.881 | 6 | 0.25 (0.10-0.51) | 0.01-0.94 | 99 | 1.67 | 0.152 |
| Unclear/ high risk | 47 | 0.19 (0.14- 0.25) | 0.02-0.71 | 86 | 1.31 | | 47 | 0.44 (0.36-0.53) | 0.04-0.94 | 96 | 2.25 | |

[a] n = number of clusters analysed, which exceeds the total number of studies
[b] Assessed in the risk of bias tool (S2 Text), with full assessments in S3 Fig;

**S5 Table. Characteristics of mathematical modelling studies and methods for estimation of the contribution of asymptomatic and presymptomatic infection to SARS-CoV-2 transmission**

| **First author, publication year [ref]** | **Original data analysed** | **Method/sources** | **Comments** | **Level of evidence** |
|---|---|---|---|---|
| Ferretti L, 2020 [1] | 40 transmission pairs, publicly available sources, China. Used to estimate generation time | Incubation period: Lauer et al. 2020 [2]<br>Serial interval estimated | Original data: "manually selected according to high confidence of direct transmission inferred from publicly available sources at the time of writing (March 2020), and with known time of onset of symptoms for both source and recipient."<br>Control measures: "effect of control measures discussed later will be relative to the early stages of an outbreak" | Moderate |
| Emery JC, 2020 [3] | Data from the Diamond Princess outbreak, 20 Jan-14 Feb 2020, extracted from Mizumoto et al. [4] Nishiura et al. [5]. | Deterministic, compartmental model.<br>Latent period: 4.3 days, duration of presymptomatic state: 2.1 days, Backer et al. [6]; duration of asymptomatic state: 5 days (assumed). | The model is fitted to two sets of data: the number symptomatic cases and the cases detected from extensive testing of individuals regardless of symptoms.<br>The model estimates the decreases in transmissibility for pre- and asymptomatic individuals. However, the wide CI in the estimate of contribution of asymptomatic infections to transmission shows an issue in identifying such parameters. | Low/moderate |
| Zhang W, 2020 [7] | No original data analysed. | Incubation period: Li et al. 2020 [8]<br>Serial interval: Li et al. 2020 [8] | Scenario 1: early transmission in Wuhan (published data from cases before 20 January 2020) | Low |
| | | Incubation period: Backer et al. 2020 [6]<br>Serial interval: Du et al. 2020 [9] | Scenario 2: Imported cases outside Wuhan (published data from cases 21 January – 8 February 2020) | |

| First author, publication year [ref] | Original data analysed | Method/sources | Comments | Level of evidence |
|---|---|---|---|---|
| | | | Control measures: "(the Wuhan lockdown was initiated on Jan. 23, 2020)." "The dramatic change of infection time distribution between these two scenarios may due to effective case isolation and quarantine of people with Wuhan travel history which could significantly reduce transmissions after symptom onset." | |
| He X, 2020 [10] | 77 transmission pairs, publicly available sources within and outside mainland China. Used to estimate serial interval. | Incubation period taken from Li et al. 2020 [8]<br>Serial interval estimated. | Original data: from several different countries in SE Asia, Europe. Dates of contact 13 January 2020 – 25 February, where reported.<br>Control measures in place at the time of data collection and heterogeneity between sources for transmission pairs not discussed. | Moderate |
| Peak CM, 2020 [11] | No original data | Estimate the infectiousness profile for two scenarios.<br>Incubation period: Li et al. 2020 [8]<br>Serial interval: Nishiura et al 2020 [5] | Scenario 1: Normal serial interval | Low |
| | | Incubation period: Li et al. 2020 [8]<br>Serial interval: Li et al. 2020 [8] | Scenario 2: Long serial interval<br>Control measures: reliability of input parameters mentioned as limitation, but not in terms of the control measures in place at the time of the original data collection | |
| Tindale LC, 2021 [12] | 54 transmission pairs in Singapore | Serial interval and incubation period estimated | They considered that incubation and serial interval are dependent.<br>Changes in estimates of incubation over time suggests the presence of intermediate cases. | High (but no CI) |
| | 80 transmission pairs in Tianjin, China | | | |

| First author, publication year [ref] | Original data analysed | Method/sources | Comments | Level of evidence |
|---|---|---|---|---|
| | | | Relatively short serial interval estimated, reflecting the contact tracing implementation.<br>Data from transmission pairs used in previous publication [13]<br>Control measures: described for each setting. Tianjin measures stronger than Singapore | |
| Moghadas SM, 2020 [14] | No original data | Incubation: Li et al. 2020 [8]<br>Presymptomatic period: Li et al. 2020 [15]<br>Infectious period from onset of symptom: He X et al. 2020 [10] | Scenario 1: Asymptomatic: 17.9% [4]<br>Scenario 2: Asymptomatic: 30.8% [5]<br>Control measures: not discussed | Low |
| Ren X, 2021 [16] | 80 transmission pairs, infectors were people who had visited Wuhan, China; infectees were outside Hubei province | For the 55 pairs where transmission could have happened before or after symptoms, Monte Carlo simulations to estimate whether the chance of asymptomatic transmission is >50% | No mathematical model, they counted the proportion of infections from presymptomatic.<br>Control measures: "We restricted the study participants to COVID-19 cases reported outside Hubei Province in the early stage of outbreaks in China, before any community transmission had occurred in these areas." | High (but no CI) |
| Chun JY, 2021 [17] | 72 transmission pairs, South Korea, until 31 March | Bayesian method to infer the infectiousness profile | Very short incubation period and serial interval estimated (2.9 and 3.6 days respectively).<br>Control measures: not discussed | High |
| Bushman M, 2021 [18] | 873 transmission pairs in China, before NPIs | Serial interval is estimated. The incubation period prior: Lauer et al. 2020 [2], Zhang et al. 2020 [19] and Backer et al. 2020 [6] | Also assumed an incubation-dependent model (incubation longer when generation interval longer).<br>The data includes some data from He X [10].<br>Control measures: "We divided case pairs into two time periods using the symptom onset | Moderate |

| First author, publication year [ref] | Original data analysed | Method/sources | Comments | Level of evidence |
|---|---|---|---|---|
| | | | dates of the primary cases. January 23 marked the lockdown of Wuhan and the start of a national rollout of nonpharmaceutical interventions (NPIs)” | |
| Sun K, 2020 [20] | Contact tracing data, Hunan China, before lockdown | The infectious profile (Fig 3E) is inferred from the transmission pairs. | Control measures: “Risk is further stratified by the date of implementation of social distancing interventions in Hunan, which is 25 January 2020.” | High (but no CI) |
| Wu P, 2021 [21] | 96 transmission pairs, contact tracing data from 4 provinces and 1 municipality in China | The authors used the same method as described by He et al. (see above). Incubation period taken from Li et al. 2020 [8].<br>Serial interval estimated. | Regarding the impact of control measure on the estimate, the authors acknowledged that “the estimate is likely to be the upper limit of contribution to the overall infections since further transmission might have been interrupted by isolation of confirmed cases depending on the efficiency in case finding” but added that their estimate “suffered little from such interruption”. | Moderate |
| Tan J, 2021 [22] | All confirmed cases with symptom status, January 7th to February 21st, 2020, Zhejiang province, China (asymptomatic or symptomatic) | Age-stratified compartmental model with distinction between presymptomatic, asymptomatic and symptomatic infections.<br>Main assumptions: Latent period of 2 days (Lauer et al. [2], Backer et al. [6]), period of presymptomatic transmission: 3-4 days (Kong et al. [23], Wei et al.) [24]. | The authors estimated the proportion of asymptomatic infections (accounting for unconfirmed asymptomatic infections) and the reduced transmissibility of asymptomatics, both by age. They could then estimate the contribution of asymptomatic to infections, overall and by age.<br>Limitation: The model involves many parameters that are estimated with only the reported number of symptomatic and asymptomatic infections, which questions about the identifiability of all the parameters | Low/moderate |

CI, confidence interval; NPI, non-pharmaceutical intervention.

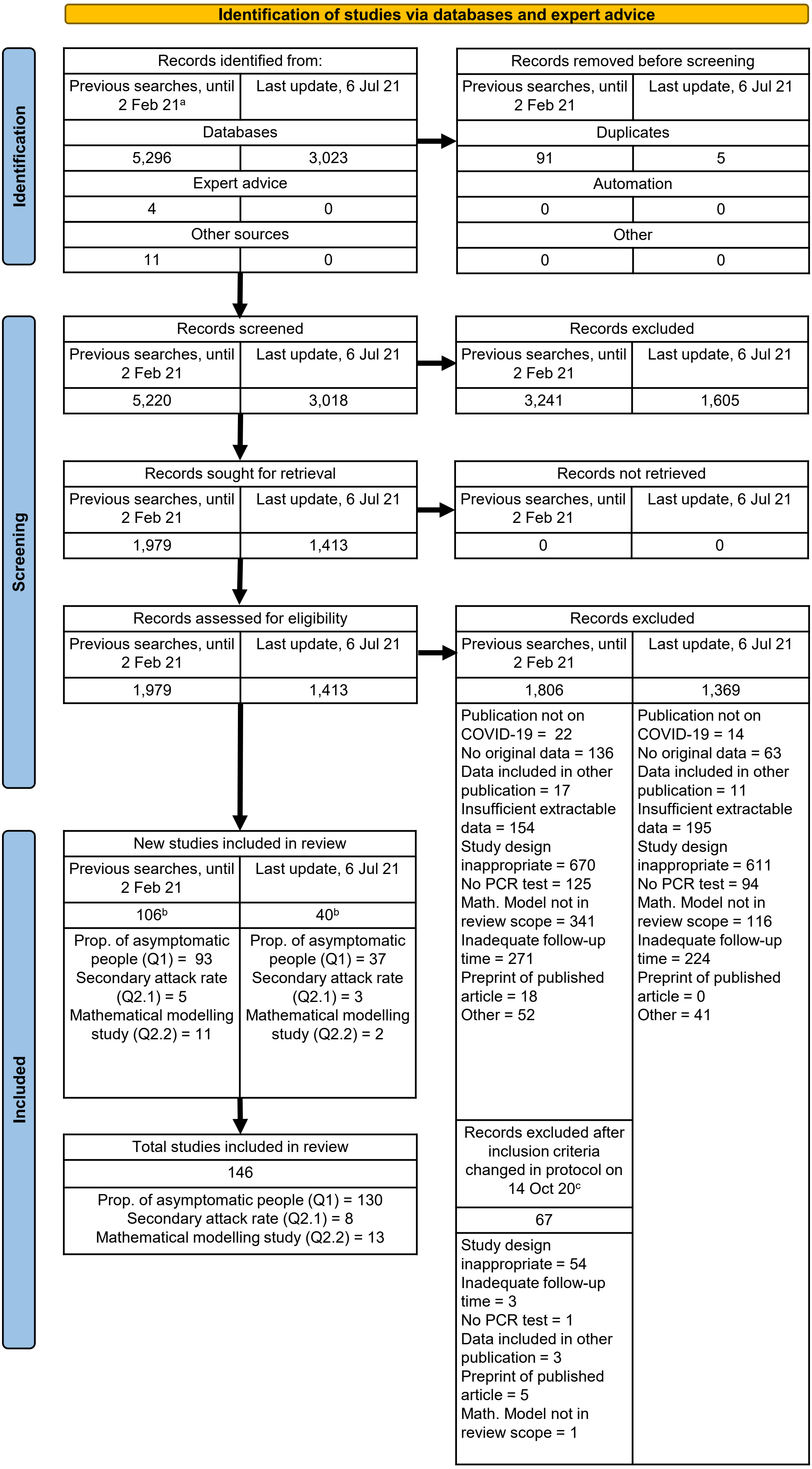

**S1 Figure. Flowchart of identified, excluded and included records as of 6 July 2021**

COVID-19, Coronavirus disease 2019; Math., mathematical; Prop., proportion;

[a]We also included papers that were published after the search date if we had previously identified their preprint.

[b]Note that some studies provided information for more than one question, therefore the total number is less than the sum of the study types.

[c]The protocol was updated on 18 Jun 21 and the study questions and inclusion criteria were updated. See S1 Table for more detail on these excluded studies.

**S2 Figure. Forest plot of proportion of people with asymptomatic SARS-CoV-2 infection, stratified by study design.**

| Study | Events | Total | Proportion | 95% CI |
|---|---|---|---|---|
| **Contact investigation** | | | | |
| Hijnen D | 1 | 11 | 0.09 | [0.00; 0.41] |
| Zhang W | 4 | 12 | 0.33 | [0.10; 0.65] |
| Cheng HY | 4 | 22 | 0.18 | [0.05; 0.40] |
| Brandstetter S | 2 | 36 | 0.06 | [0.01; 0.19] |
| Yousaf AR | 0 | 47 | 0.00 | [0.00; 0.08] |
| Wu J | 5 | 48 | 0.10 | [0.03; 0.23] |
| Xie W | 4 | 53 | 0.08 | [0.02; 0.18] |
| Gettings JR | 31 | 55 | 0.56 | [0.42; 0.70] |
| Jones A | 24 | 87 | 0.28 | [0.19; 0.38] |
| Grijalva CG | 34 | 102 | 0.33 | [0.24; 0.43] |
| Luo L2 | 8 | 127 | 0.06 | [0.03; 0.12] |
| Shi Q | 60 | 183 | 0.33 | [0.26; 0.40] |
| Hurst JH | 87 | 293 | 0.30 | [0.25; 0.35] |
| **Prediction interval** | . | . | | **[0.01; 0.81]** |
| **Outbreak investigation** | | | | |
| Pirnay JP | 2 | 4 | 0.50 | [0.07; 0.93] |
| Cardillo L [Healthcare workers] | 2 | 4 | 0.50 | [0.07; 0.93] |
| Garibaldi [Staff] | 0 | 8 | 0.00 | [0.00; 0.37] |
| Corcorran MA | 3 | 8 | 0.38 | [0.09; 0.76] |
| Yang N | 2 | 10 | 0.20 | [0.03; 0.56] |
| Schwierzeck V | 2 | 11 | 0.18 | [0.02; 0.52] |
| Danis K | 1 | 12 | 0.08 | [0.00; 0.38] |
| Garibaldi [Residents] | 1 | 12 | 0.08 | [0.00; 0.38] |
| Romao VC | 0 | 14 | 0.00 | [0.00; 0.23] |
| Orsi A | 13 | 14 | 0.93 | [0.66; 1.00] |
| Böhmer MM | 1 | 16 | 0.06 | [0.00; 0.30] |
| Dora AV | 6 | 16 | 0.38 | [0.15; 0.65] |
| Yau K | 7 | 20 | 0.35 | [0.15; 0.59] |
| Redditt V | 3 | 24 | 0.12 | [0.03; 0.32] |
| Tian S | 7 | 24 | 0.29 | [0.13; 0.51] |
| Harada S [Patients] | 8 | 24 | 0.33 | [0.16; 0.55] |
| Park JH | 4 | 28 | 0.14 | [0.04; 0.33] |
| Patel MC | 13 | 35 | 0.37 | [0.21; 0.55] |
| Kittang BR | 0 | 40 | 0.00 | [0.00; 0.09] |
| Pavli A | 7 | 46 | 0.15 | [0.06; 0.29] |
| Arons MM | 3 | 47 | 0.06 | [0.01; 0.18] |
| Harada S [Healthcare workers] | 25 | 49 | 0.51 | [0.36; 0.66] |
| Ladhani SN [Healthcare workers] | 26 | 53 | 0.49 | [0.35; 0.63] |
| van den Besselaar JH [Healthcare workers] | 1 | 54 | 0.02 | [0.00; 0.10] |
| Schmitt J | 18 | 54 | 0.33 | [0.21; 0.47] |
| Plucinski MM | 14 | 66 | 0.21 | [0.12; 0.33] |
| Njuguna H | 29 | 71 | 0.41 | [0.29; 0.53] |
| Cardillo L [Patients] | 20 | 91 | 0.22 | [0.14; 0.32] |
| Park SY | 4 | 95 | 0.04 | [0.01; 0.10] |
| Taylor J [Healthcare personnel] | 9 | 99 | 0.09 | [0.04; 0.17] |
| Ladhani SN [Residents] | 46 | 105 | 0.44 | [0.34; 0.54] |
| Paleker M | 41 | 112 | 0.37 | [0.28; 0.46] |
| van den Besselaar JH [Residents] | 7 | 113 | 0.06 | [0.03; 0.12] |
| Graham N | 46 | 126 | 0.37 | [0.28; 0.46] |
| Taylor J [Residents] | 7 | 127 | 0.06 | [0.02; 0.11] |
| Pham QT | 89 | 208 | 0.43 | [0.36; 0.50] |
| Kennelly SP [Nursing home staff] | 97 | 395 | 0.25 | [0.20; 0.29] |
| Lee JY | 80 | 694 | 0.12 | [0.09; 0.14] |
| Kennelly SP [Nursing home residents] | 193 | 710 | 0.27 | [0.24; 0.31] |
| Kasper MR | 572 | 1271 | 0.45 | [0.42; 0.48] |
| **Prediction interval** | . | . | | **[0.01; 0.81]** |
| **Screening: institutional setting** | | | | |
| Tanacan A | 0 | 3 | 0.00 | [0.00; 0.71] |
| Jeffery−Smith A [Staff] | 2 | 3 | 0.67 | [0.09; 0.99] |
| Fisher MJ | 3 | 4 | 0.75 | [0.19; 0.99] |
| Berghoff AS | 2 | 4 | 0.50 | [0.07; 0.93] |
| Vohra [Undergoing chemotherapy] | 2 | 4 | 0.50 | [0.07; 0.93] |
| Pamplona J | 2 | 5 | 0.40 | [0.05; 0.85] |
| Viñuela MC | 8 | 8 | 1.00 | [0.63; 1.00] |
| Bender WR | 6 | 8 | 0.75 | [0.35; 0.97] |
| van Buul LW [Healthcare workers] | 0 | 9 | 0.00 | [0.00; 0.34] |
| Vohra [Presurgical patients] | 10 | 10 | 1.00 | [0.69; 1.00] |
| Varnell C | 5 | 10 | 0.50 | [0.19; 0.81] |
| Haidar G | 9 | 11 | 0.82 | [0.48; 0.98] |
| Kirshblum SC | 2 | 12 | 0.17 | [0.02; 0.48] |
| Jeffery−Smith A [Residents] | 6 | 13 | 0.46 | [0.19; 0.75] |
| Isoldi S | 4 | 15 | 0.27 | [0.08; 0.55] |
| van Buul LW [Nursing home residents] | 3 | 16 | 0.19 | [0.04; 0.46] |
| Wadhwa A | 6 | 17 | 0.35 | [0.14; 0.62] |
| Balestrini S | 11 | 17 | 0.65 | [0.38; 0.86] |
| Maki G | 16 | 18 | 0.89 | [0.65; 0.99] |
| Bogani G | 10 | 19 | 0.53 | [0.29; 0.76] |
| Green R | 22 | 22 | 1.00 | [0.85; 1.00] |
| Martins Machado C | 5 | 22 | 0.23 | [0.08; 0.45] |
| Khondaker T | 7 | 26 | 0.27 | [0.12; 0.48] |
| Starling A | 29 | 31 | 0.94 | [0.79; 0.99] |
| Rincon A | 9 | 35 | 0.26 | [0.12; 0.43] |
| Pizarro−Sánchez MS | 5 | 38 | 0.13 | [0.04; 0.28] |
| Patel MR | 43 | 55 | 0.78 | [0.65; 0.88] |
| Aslam A | 38 | 65 | 0.58 | [0.46; 0.71] |
| Marossy A | 46 | 67 | 0.69 | [0.56; 0.79] |
| London V | 22 | 68 | 0.32 | [0.22; 0.45] |
| Smith E | 42 | 103 | 0.41 | [0.31; 0.51] |
| Weinbergerova B | 6 | 105 | 0.06 | [0.02; 0.12] |
| Esteban I | 75 | 113 | 0.66 | [0.57; 0.75] |
| Nunes MC | 14 | 115 | 0.12 | [0.07; 0.20] |
| Hcini N | 87 | 137 | 0.64 | [0.55; 0.72] |
| Shi SM | 21 | 146 | 0.14 | [0.09; 0.21] |
| Andrikopoulou M | 46 | 158 | 0.29 | [0.22; 0.37] |
| Blain H | 14 | 161 | 0.09 | [0.05; 0.14] |
| Say D | 61 | 171 | 0.36 | [0.29; 0.43] |
| Beiting KJ | 50 | 172 | 0.29 | [0.22; 0.36] |
| Uçkay, I | 71 | 175 | 0.41 | [0.33; 0.48] |
| Adhikari EH | 98 | 252 | 0.39 | [0.33; 0.45] |
| Hussain A | 54 | 266 | 0.20 | [0.16; 0.26] |
| Marcus JE | 199 | 403 | 0.49 | [0.44; 0.54] |
| Ghinai I | 293 | 406 | 0.72 | [0.68; 0.76] |
| White EM | 2194 | 5403 | 0.41 | [0.39; 0.42] |
| **Prediction interval** | . | . | | **[0.05; 0.93]** |
| **Screening: community setting** | | | | |
| Hoehl S | 1 | 2 | 0.50 | [0.01; 0.99] |
| Chang L | 2 | 4 | 0.50 | [0.07; 0.93] |
| Rauch JN | 2 | 6 | 0.33 | [0.04; 0.78] |
| AbdulRahman A | 3 | 6 | 0.50 | [0.12; 0.88] |
| Theuring S [School students and staff] | 2 | 10 | 0.20 | [0.03; 0.56] |
| Kutsuna S | 3 | 11 | 0.27 | [0.06; 0.61] |
| Theuring S [Household members] | 0 | 14 | 0.00 | [0.00; 0.23] |
| Hwang | 8 | 14 | 0.57 | [0.29; 0.82] |
| Edelstein M | 4 | 20 | 0.20 | [0.06; 0.44] |
| Migisha R | 20 | 54 | 0.37 | [0.24; 0.51] |
| Lavezzo E | 29 | 73 | 0.40 | [0.28; 0.52] |
| Chamie G | 23 | 81 | 0.28 | [0.19; 0.40] |
| Meyers KJ | 67 | 86 | 0.78 | [0.68; 0.86] |
| Wi YM | 7 | 111 | 0.06 | [0.03; 0.13] |
| Turunen T | 23 | 127 | 0.18 | [0.12; 0.26] |
| Wong J | 16 | 138 | 0.12 | [0.07; 0.18] |
| Eythorsson E | 25 | 178 | 0.14 | [0.09; 0.20] |
| Al−Qahtani M | 116 | 188 | 0.62 | [0.54; 0.69] |
| Cao S | 300 | 300 | 1.00 | [0.99; 1.00] |
| Uysal E | 64 | 684 | 0.09 | [0.07; 0.12] |
| Almazeedi S | 473 | 1096 | 0.43 | [0.40; 0.46] |
| Malhotra S | 1272 | 1729 | 0.74 | [0.71; 0.76] |
| Abraha HE | 1935 | 2617 | 0.74 | [0.72; 0.76] |
| Ren R | 1612 | 3103 | 0.52 | [0.50; 0.54] |
| **Prediction interval** | . | . | | **[0.03; 0.92]** |
| **Screening: occupational** | | | | |
| Lalani T | 4 | 8 | 0.50 | [0.16; 0.84] |
| Ferreira VH | 5 | 9 | 0.56 | [0.21; 0.86] |
| Morgan SC | 2 | 11 | 0.18 | [0.02; 0.52] |
| Han X | 17 | 17 | 1.00 | [0.80; 1.00] |
| Alshahrani MS | 12 | 18 | 0.67 | [0.41; 0.87] |
| Stock AD | 6 | 19 | 0.32 | [0.13; 0.57] |
| Fakhim H | 14 | 21 | 0.67 | [0.43; 0.85] |
| Laws RL | 25 | 28 | 0.89 | [0.72; 0.98] |
| Rivett L | 5 | 30 | 0.17 | [0.06; 0.35] |
| Malagón−Rojas J | 11 | 35 | 0.31 | [0.17; 0.49] |
| Hogan CA | 20 | 38 | 0.53 | [0.36; 0.69] |
| Treibel TA | 12 | 44 | 0.27 | [0.15; 0.43] |
| Tan−Loh J | 6 | 46 | 0.13 | [0.05; 0.26] |
| Letizia AG | 46 | 51 | 0.90 | [0.79; 0.97] |
| Lombardi A | 17 | 139 | 0.12 | [0.07; 0.19] |
| Cariani L | 32 | 182 | 0.18 | [0.12; 0.24] |
| Mahajan NN | 58 | 467 | 0.12 | [0.10; 0.16] |
| Hall VJ | 293 | 1704 | 0.17 | [0.15; 0.19] |
| **Prediction interval** | . | . | | **[0.03; 0.93]** |
| **Prediction interval** | | | | **[0.02; 0.90]** |

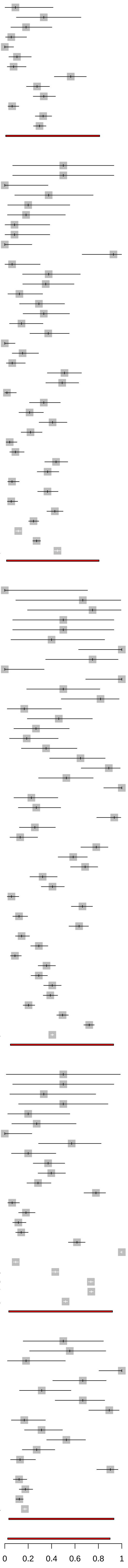

**S3 Fig. Risk of bias assessment of studies in review question 1 and 2**

### Question 1

Contact investigation

| Study | Risk of bias items 1 | 2 | 3 | 4 | 5 | 6 |
|---|---|---|---|---|---|---|
| Cheng HY | + | + | X | - | - | + |
| Wu J | X | - | + | - | + | - |
| Hijnen D | + | - | - | - | - | + |
| Brandstetter S | - | - | + | - | + | + |
| Zhang W | + | + | - | - | + | + |
| Yousaf AR | + | - | + | + | + | + |
| Luo L2 | + | + | + | + | + | + |
| Hurst JH | X | + | + | X | X | X |
| Grijalva CG | - | - | - | + | X | + |
| Shi Q | + | + | - | - | + | + |
| Jones A | - | - | - | - | + | + |
| Xie W | + | + | + | - | + | + |
| Gettings JR | - | - | + | + | X | X |

Outbreak investigation

Risk of bias items

| Study | 1 | 2 | 3 | 4 | 5 | 6 |
|---|---|---|---|---|---|---|
| Danis K | - | - | + | + | + | + |
| Park SY | + | + | - | - | + | + |
| Arons MM | + | + | + | + | + | + |
| Schwierzeck V | + | + | - | + | + | + |
| Böhmer MM | + | + | + | - | + | + |
| Dora AV | + | + | X | + | + | + |
| Graham N | + | + | X | X | X | + |
| Patel MC | + | + | + | + | + | + |
| Pavli A | + | + | + | - | + | + |
| Njuguna H | - | + | X | X | + | + |
| Yau K | + | + | - | - | + | + |
| Lee JY | - | - | X | + | X | + |
| Yang N | + | + | + | + | + | + |
| Pham QT | + | + | - | - | + | + |
| Plucinski MM | X | X | + | X | X | + |
| Kittang BR | + | - | + | - | + | + |
| Corcoran MA | + | + | + | + | + | + |
| Pirnay JP | + | + | + | - | + | + |
| Taylor J | - | - | - | - | + | + |
| Kennelly SP | + | - | X | - | X | + |
| Romao VC | + | - | + | + | + | + |
| Ladhani SN | + | + | - | - | + | + |
| Kasper MR | + | + | + | + | + | + |
| Redditt V | + | - | + | X | - | + |
| Park JH | + | + | X | - | + | + |
| Harada S | + | - | + | + | + | + |
| Garibaldi PMM | + | + | - | + | X | + |
| Tian S | + | + | + | - | + | + |
| van den Besselaar JH | + | X | + | - | + | X |
| Orsi A | + | + | - | - | + | + |
| Schmitt J | + | + | - | + | - | + |
| Paleker M | - | - | - | - | + | X |
| Cardillo L | - | - | X | - | + | + |

Screening: community setting

| Study | Risk of bias items 1 | 2 | 3 | 4 | 5 | 6 |
|---|---|---|---|---|---|---|
| Chang L | X | + | - | - | + | + |
| Lavezzo E | + | - | + | X | + | + |
| Hoehl S | + | + | + | + | + | + |
| Wong J | + | - | - | - | + | + |
| Almazeedi S | + | - | + | - | + | + |
| Wi YM | + | + | + | + | + | + |
| Kutsuna S | + | - | - | + | + | + |
| Eythorsson E | + | - | + | + | X | + |
| Chamie G | X | - | - | - | + | X |
| AbdulRahman A | X | - | - | - | + | + |
| Edelstein M | X | - | X | X | + | + |
| Al-Qahtani M | + | + | X | + | + | + |
| Cao S | + | - | X | - | + | + |
| Migisha R | - | - | + | + | - | + |
| Meyers KJ | X | - | + | X | + | + |
| Ren R | + | + | - | + | + | + |
| Rauch JN | X | - | - | - | X | + |
| Uysal E | + | + | + | - | + | + |
| Abraha HE | + | + | + | + | + | + |
| Turunen T | + | - | + | + | + | + |
| Malhotra S | X | - | X | - | X | + |
| Hwang | + | + | + | + | - | + |
| Theuring S | - | - | X | + | X | + |

Screening: institutional setting

| Study | Risk of bias items 1 | 2 | 3 | 4 | 5 | 6 |
|---|---|---|---|---|---|---|
| London V | + | + | X | - | + | + |
| Andrikopoulou M | + | + | + | + | + | + |
| Bogani G | + | + | X | + | + | + |
| Kirshblum SC | + | + | X | + | + | + |
| Smith E | + | - | + | - | + | + |
| Tanacan A | X | - | - | - | + | + |
| Starling A | - | - | X | - | + | + |
| Marossy A | + | - | - | - | + | + |
| Viñuela MC | + | + | - | - | + | + |
| Bender WR | X | X | + | X | - | + |
| Shi SM | X | X | - | + | + | + |
| Berghoff AS | + | + | - | - | + | + |
| White EM | - | + | + | + | - | + |
| Aslam A | + | + | - | + | + | + |
| Adhikari EH | X | - | - | - | - | + |
| Ghinai I | + | - | + | X | + | X |
| Wadhwa A | + | X | + | X | X | + |
| Balestrini S | + | + | - | - | + | + |
| Hcini N | + | + | - | - | + | + |
| van Buul LW | + | - | + | - | + | + |
| Green R | + | - | - | + | - | + |
| Varnell C | X | - | + | - | + | - |
| Pizarro-Sánchez MS | X | X | - | - | + | X |
| Vohra LM | + | + | X | - | + | + |
| Isoldi S | + | - | + | - | + | X |
| Haidar G | X | X | + | + | + | + |
| Blain H | + | + | + | + | + | + |
| Marcus JE | + | + | + | - | + | + |
| Rincon A | + | + | + | + | + | + |
| Martins Machado C | + | - | - | - | + | + |
| Beiting KJ | + | - | + | + | + | + |
| Hussain A | X | - | + | - | + | + |
| Maki G | X | + | - | - | + | + |
| Pamplona J | X | - | X | - | + | + |
| Jeffery-Smith A | - | - | + | + | + | + |
| Uçkay, I | - | - | - | - | X | + |
| Say D | - | - | + | - | + | + |
| Nunes MC | - | - | + | + | + | + |
| Weinbergerova B | X | - | + | + | + | + |
| Esteban I | X | - | + | + | + | + |
| Fisher MJ | + | - | X | - | X | - |
| Patel MR | + | + | X | - | + | + |
| Khondaker T | + | + | X | - | + | + |

Screening: occupational

| Study | Risk of bias items: 1 | 2 | 3 | 4 | 5 | 6 |
|---|---|---|---|---|---|---|
| Rivett L | X | X | + | + | - | + |
| Treibel TA | - | - | - | - | X | + |
| Han X | X | - | - | - | X | + |
| Lombardi A | + | + | + | - | + | + |
| Cariani L | + | + | - | - | + | + |
| Stock AD | - | - | - | - | + | + |
| Malagón-Rojas J | - | - | - | X | + | + |
| Letizia AG | X | - | - | - | + | + |
| Mahajan NN | - | - | X | + | + | X |
| Alshahrani MS | + | + | + | X | + | + |
| Hogan CA | X | - | + | - | X | + |
| Ferreira VH | X | - | + | + | - | + |
| Tan-Loh J | - | - | + | + | + | X |
| Lalani T | + | - | X | X | X | X |
| Fakhim H | X | - | - | - | + | + |
| Hall VJ | X | - | + | X | + | + |
| Laws RL | + | + | - | - | + | + |
| Morgan SC | X | - | - | - | + | + |

Judgement

+ Low
- Unclear
X High

Domains

Selection Bias:
1: Representativeness of the sample
2: Characteristics of non-respondents
Information Bias:
3: Symptom assessment
4: Recording of symptoms
Misclassification bias:
5: Classification of asymptomatic status
Attrition bias:
6: Selective reporting of symptoms status

### Question 2.1

Risk of bias items

| Study | 1 | 2 | 3 | 4 | 5 | 6 |
|---|---|---|---|---|---|---|
| Luo L1 | + | + | - | + | + | + |
| Park SY | + | + | - | - | + | + |
| Cheng HY | + | + | X | - | - | + |
| Zhang W | + | + | - | - | + | + |
| Chaw L | - | - | X | + | + | + |
| Bender JK | + | - | + | + | + | + |
| Wu P | + | + | + | - | - | + |
| Gettings JR | - | - | + | + | X | X |

**Judgement**

- + Low
- - Unclear
- X High

Domains

Selection Bias:
1: Representativeness of the sample
2: Characteristics of non-respondents
Information Bias:
3: Symptom assessment
4: Recording of symptoms
Misclassification bias:
5: Classification of asymptomatic status
Attrition bias:
6: Selective reporting of symptoms status

| Study | Events | Total | Proportion | 95% CI |
|---|---|---|---|---|
| **Published Jan '20 − Jun '20** | | | | |
| Hoehl S | 1 | 2 | 0.50 | [0.01; 0.99] |
| Chang L | 2 | 4 | 0.50 | [0.07; 0.93] |
| Hijnen D | 1 | 11 | 0.09 | [0.00; 0.41] |
| Schwierzeck V | 2 | 11 | 0.18 | [0.02; 0.52] |
| Danis K | 1 | 12 | 0.08 | [0.00; 0.38] |
| Zhang W | 4 | 12 | 0.33 | [0.10; 0.65] |
| Böhmer MM | 1 | 16 | 0.06 | [0.00; 0.30] |
| Dora AV | 6 | 16 | 0.38 | [0.15; 0.65] |
| Han X | 17 | 17 | 1.00 | [0.80; 1.00] |
| Bogani G | 10 | 19 | 0.53 | [0.29; 0.76] |
| Cheng HY | 4 | 22 | 0.18 | [0.05; 0.40] |
| Rivett L | 5 | 30 | 0.17 | [0.06; 0.35] |
| Patel MC | 13 | 35 | 0.37 | [0.21; 0.55] |
| Brandstetter S | 2 | 36 | 0.06 | [0.01; 0.19] |
| Treibel TA | 12 | 44 | 0.27 | [0.15; 0.43] |
| Pavli A | 7 | 46 | 0.15 | [0.06; 0.29] |
| Arons MM | 3 | 47 | 0.06 | [0.01; 0.18] |
| Wu J | 5 | 48 | 0.10 | [0.03; 0.23] |
| London V | 22 | 68 | 0.32 | [0.22; 0.45] |
| Park SY | 4 | 95 | 0.04 | [0.01; 0.10] |
| Graham N | 46 | 126 | 0.37 | [0.28; 0.46] |
| Wong J | 16 | 138 | 0.12 | [0.07; 0.18] |
| Lombardi A | 17 | 139 | 0.12 | [0.07; 0.19] |
| Andrikopoulou M | 46 | 158 | 0.29 | [0.22; 0.37] |
| **Prediction interval** | . | . | | **[0.01; 0.85]** |
| **Published Jul '20 − Dec '20** | | | | |
| Tanacan A | 0 | 3 | 0.00 | [0.00; 0.71] |
| Pirnay JP | 2 | 4 | 0.50 | [0.07; 0.93] |
| Berghoff AS | 2 | 4 | 0.50 | [0.07; 0.93] |
| AbdulRahman A | 3 | 6 | 0.50 | [0.12; 0.88] |
| Corcorran MA | 3 | 8 | 0.38 | [0.09; 0.76] |
| Viñuela MC | 8 | 8 | 1.00 | [0.63; 1.00] |
| Bender WR | 6 | 8 | 0.75 | [0.35; 0.97] |
| Yang N | 2 | 10 | 0.20 | [0.03; 0.56] |
| Kutsuna S | 3 | 11 | 0.27 | [0.06; 0.61] |
| Kirshblum SC | 2 | 12 | 0.17 | [0.02; 0.48] |
| Romao VC | 0 | 14 | 0.00 | [0.00; 0.23] |
| Wadhwa A | 6 | 17 | 0.35 | [0.14; 0.62] |
| Balestrini S | 11 | 17 | 0.65 | [0.38; 0.86] |
| Alshahrani MS | 12 | 18 | 0.67 | [0.41; 0.87] |
| Stock AD | 6 | 19 | 0.32 | [0.13; 0.57] |
| Yau K | 7 | 20 | 0.35 | [0.15; 0.59] |
| Edelstein M | 4 | 20 | 0.20 | [0.06; 0.44] |
| Redditt V | 3 | 24 | 0.12 | [0.03; 0.32] |
| Harada S [Patients] | 8 | 24 | 0.33 | [0.16; 0.55] |
| Park JH | 4 | 28 | 0.14 | [0.04; 0.33] |
| Starling A | 29 | 31 | 0.94 | [0.79; 0.99] |
| Malagón−Rojas J | 11 | 35 | 0.31 | [0.17; 0.49] |
| Kittang BR | 0 | 40 | 0.00 | [0.00; 0.09] |
| Yousaf AR | 0 | 47 | 0.00 | [0.00; 0.08] |
| Harada S [Healthcare workers] | 25 | 49 | 0.51 | [0.36; 0.66] |
| Letizia AG | 46 | 51 | 0.90 | [0.79; 0.97] |
| Ladhani SN [Healthcare workers] | 26 | 53 | 0.49 | [0.35; 0.63] |
| Migisha R | 20 | 54 | 0.37 | [0.24; 0.51] |
| Aslam A | 38 | 65 | 0.58 | [0.46; 0.71] |
| Plucinski MM | 14 | 66 | 0.21 | [0.12; 0.33] |
| Marossy A | 46 | 67 | 0.69 | [0.56; 0.79] |
| Njuguna H | 29 | 71 | 0.41 | [0.29; 0.53] |
| Lavezzo E | 29 | 73 | 0.40 | [0.28; 0.52] |
| Chamie G | 23 | 81 | 0.28 | [0.19; 0.40] |
| Taylor J [Healthcare personnel] | 9 | 99 | 0.09 | [0.04; 0.17] |
| Grijalva CG | 34 | 102 | 0.33 | [0.24; 0.43] |
| Smith E | 42 | 103 | 0.41 | [0.31; 0.51] |
| Ladhani SN [Residents] | 46 | 105 | 0.44 | [0.34; 0.54] |
| Wi YM | 7 | 111 | 0.06 | [0.03; 0.13] |
| Taylor J [Residents] | 7 | 127 | 0.06 | [0.02; 0.11] |
| Luo L2 | 8 | 127 | 0.06 | [0.03; 0.12] |
| Hcini N | 87 | 137 | 0.64 | [0.55; 0.72] |
| Shi SM | 21 | 146 | 0.14 | [0.09; 0.21] |
| Eythorsson E | 25 | 178 | 0.14 | [0.09; 0.20] |
| Cariani L | 32 | 182 | 0.18 | [0.12; 0.24] |
| Shi Q | 60 | 183 | 0.33 | [0.26; 0.40] |
| Al−Qahtani M | 116 | 188 | 0.62 | [0.54; 0.69] |
| Pham QT | 89 | 208 | 0.43 | [0.36; 0.50] |
| Adhikari EH | 98 | 252 | 0.39 | [0.33; 0.45] |
| Hurst JH | 87 | 293 | 0.30 | [0.25; 0.35] |
| Cao S | 300 | 300 | 1.00 | [0.99; 1.00] |
| Kennelly SP [Nursing home staff] | 97 | 395 | 0.25 | [0.20; 0.29] |
| Ghinai I | 293 | 406 | 0.72 | [0.68; 0.76] |
| Mahajan NN | 58 | 467 | 0.12 | [0.10; 0.16] |
| Lee JY | 80 | 694 | 0.12 | [0.09; 0.14] |
| Kennelly SP [Nursing home residents] | 193 | 710 | 0.27 | [0.24; 0.31] |
| Almazeedi S | 473 | 1096 | 0.43 | [0.40; 0.46] |
| Kasper MR | 572 | 1271 | 0.45 | [0.42; 0.48] |
| White EM | 2194 | 5403 | 0.41 | [0.39; 0.42] |
| **Prediction interval** | . | . | | **[0.03; 0.91]** |
| **Published from Jan '21 onwards** | | | | |
| Jeffery−Smith A [Staff] | 2 | 3 | 0.67 | [0.09; 0.99] |
| Cardillo L [Healthcare workers] | 2 | 4 | 0.50 | [0.07; 0.93] |
| Fisher MJ | 3 | 4 | 0.75 | [0.19; 0.99] |
| Vohra [Undergoing chemotherapy] | 2 | 4 | 0.50 | [0.07; 0.93] |
| Pamplona J | 2 | 5 | 0.40 | [0.05; 0.85] |
| Rauch JN | 2 | 6 | 0.33 | [0.04; 0.78] |
| Garibaldi [Staff] | 0 | 8 | 0.00 | [0.00; 0.37] |
| Lalani T | 4 | 8 | 0.50 | [0.16; 0.84] |
| van Buul LW [Healthcare workers] | 0 | 9 | 0.00 | [0.00; 0.34] |
| Ferreira VH | 5 | 9 | 0.56 | [0.21; 0.86] |
| Vohra [Presurgical patients] | 10 | 10 | 1.00 | [0.69; 1.00] |
| Theuring S [School students and staff] | 2 | 10 | 0.20 | [0.03; 0.56] |
| Varnell C | 5 | 10 | 0.50 | [0.19; 0.81] |
| Haidar G | 9 | 11 | 0.82 | [0.48; 0.98] |
| Morgan SC | 2 | 11 | 0.18 | [0.02; 0.52] |
| Garibaldi [Residents] | 1 | 12 | 0.08 | [0.00; 0.38] |
| Jeffery−Smith A [Residents] | 6 | 13 | 0.46 | [0.19; 0.75] |
| Orsi A | 13 | 14 | 0.93 | [0.66; 1.00] |
| Theuring S [Household members] | 0 | 14 | 0.00 | [0.00; 0.23] |
| Hwang | 8 | 14 | 0.57 | [0.29; 0.82] |
| Isoldi S | 4 | 15 | 0.27 | [0.08; 0.55] |
| van Buul LW [Nursing home residents] | 3 | 16 | 0.19 | [0.04; 0.46] |
| Maki G | 16 | 18 | 0.89 | [0.65; 0.99] |
| Fakhim H | 14 | 21 | 0.67 | [0.43; 0.85] |
| Green R | 22 | 22 | 1.00 | [0.85; 1.00] |
| Martins Machado C | 5 | 22 | 0.23 | [0.08; 0.45] |
| Tian S | 7 | 24 | 0.29 | [0.13; 0.51] |
| Khondaker T | 7 | 26 | 0.27 | [0.12; 0.48] |
| Laws RL | 25 | 28 | 0.89 | [0.72; 0.98] |
| Rincon A | 9 | 35 | 0.26 | [0.12; 0.43] |
| Pizarro−Sánchez MS | 5 | 38 | 0.13 | [0.04; 0.28] |
| Hogan CA | 20 | 38 | 0.53 | [0.36; 0.69] |
| Tan−Loh J | 6 | 46 | 0.13 | [0.05; 0.26] |
| Xie W | 4 | 53 | 0.08 | [0.02; 0.18] |
| van den Besselaar JH [Healthcare workers] | 1 | 54 | 0.02 | [0.00; 0.10] |
| Schmitt J | 18 | 54 | 0.33 | [0.21; 0.47] |
| Gettings JR | 31 | 55 | 0.56 | [0.42; 0.70] |
| Patel MR | 43 | 55 | 0.78 | [0.65; 0.88] |
| Meyers KJ | 67 | 86 | 0.78 | [0.68; 0.86] |
| Jones A | 24 | 87 | 0.28 | [0.19; 0.38] |
| Cardillo L [Patients] | 20 | 91 | 0.22 | [0.14; 0.32] |
| Weinbergerova B | 6 | 105 | 0.06 | [0.02; 0.12] |
| Paleker M | 41 | 112 | 0.37 | [0.28; 0.46] |
| van den Besselaar JH [Residents] | 7 | 113 | 0.06 | [0.03; 0.12] |
| Esteban I | 75 | 113 | 0.66 | [0.57; 0.75] |
| Nunes MC | 14 | 115 | 0.12 | [0.07; 0.20] |
| Turunen T | 23 | 127 | 0.18 | [0.12; 0.26] |
| Blain H | 14 | 161 | 0.09 | [0.05; 0.14] |
| Say D | 61 | 171 | 0.36 | [0.29; 0.43] |
| Beiting KJ | 50 | 172 | 0.29 | [0.22; 0.36] |
| Uçkay, I | 71 | 175 | 0.41 | [0.33; 0.48] |
| Hussain A | 54 | 266 | 0.20 | [0.16; 0.26] |
| Marcus JE | 199 | 403 | 0.49 | [0.44; 0.54] |
| Uysal E | 64 | 684 | 0.09 | [0.07; 0.12] |
| Hall VJ | 293 | 1704 | 0.17 | [0.15; 0.19] |
| Malhotra S | 1272 | 1729 | 0.74 | [0.71; 0.76] |
| Abraha HE | 1935 | 2617 | 0.74 | [0.72; 0.76] |
| Ren R | 1612 | 3103 | 0.52 | [0.50; 0.54] |
| **Prediction interval** | . | . | | **[0.03; 0.91]** |
| **Prediction interval** | | | | **[0.02; 0.90]** |

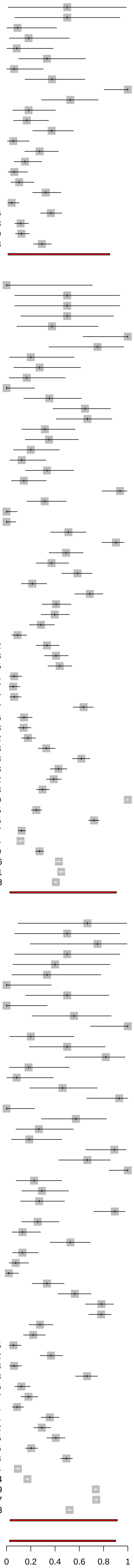

#### S5 Figure. Assessment of credibility of mathematical modelling studies.

| AUTHOR[a] | Is the population relevant? | Are any critical interventions missing? | Is the context (settings and circumstances) applicable? | Is external validation of the model sufficient to make its results credible for your decision? | Is internal verification of the model sufficient to make its results credible for your decision? | Does the model have sufficient face validity to make its results credible for your decision? | Is the design of the model adequate for your decision problem? | Are the data used in populating the model suitable for your decision problem? | Were the analyses performed using the model adequate to inform your decision problem? | Was there an adequate assessment of the effects of uncertainty? | Was the reporting of the model adequate to inform your decision problem? | Was the interpretation of results fair and balanced? | Were there any potential conflicts of interest? |
|---|---|---|---|---|---|---|---|---|---|---|---|---|---|
| **Ferreti, L** | PY | NA | YES | NO | YES | YES | YES | PY | YES | YES | YES | NO | NO |
| **Zhang, W** | PY | NA | YES | NO | YES | NO | PY | PY | NO | NO | NO | NO | NR |
| **He, X** | PY | NA | YES | NO | YES | YES | YES | PY | YES | YES | PY | YES | NO |
| **Casey, M** | YES | NA | YES | PY | YES | PY | YES | YES | PY | YES | YES | YES | NO |
| **Peak, CM** | YES | NA | PY | NO | YES | PY | YES | NO | PY | PY | PY | YES | NO |
| **Emery, JC** | YES | NA | PY | YES | YES | YES | YES | PY | YES | YES | YES | YES | NO |
| **Tindale LC** | YES | NA | YES | YES | YES | YES | PY | YES | YES | YES | YES | YES | NO |
| **Moghadas, SM** | YES | NA | YES | NO | NO | NO | YES | YES | YES | PY | NO | NO | NO |
| **Ren, X** | YES | NA | YES | NO | NO | NO | NO | YES | YES | NO | YES | PY | NO |
| **Chun, JY** | YES | NA | YES | YES | YES | YES | YES | YES | YES | YES | YES | YES | NO |
| **Sun, K** | YES | NA | YES | PY | PY | NEI | PY | YES | YES | PY | YES | YES | NO |
| **Bushman, M** | YES | NA | YES | NO | YES | NO | YES | YES | YES | YES | YES | YES | NO |
| **Wu, P** | YES | NA | YES | YES | YES | YES | YES | YES | YES | YES | YES | YES | NO |
| **Tan, J** | YES | NA | YES | NO | NO | YES | YES | YES | YES | YES | YES | YES | NO |

a. Reference numbers as per the main text

PY: Partially yes

NA: Not applicable

NR: Not reported

NEI: Not enough Information

### S1 Appendix. Data extraction forms

#### Screening Form

Record ID:

Screening assigned to [screen1_assigned]; performed by:
(change only if assignment has changed)

Verification assigned to [screen1_veri]; performed by:
(change only if assignment has changed)

Resolution assigned to [screen1_resolve]; performed by:
(change only if assignment has changed)

record_no [references redcap database]

preprint refnr [references redcap database]

PDF of article

##### Citation meta data

Authors

First author
(automatically imported; Lastname, IN)

Publication year

Title

Abstract

Author 1

Journal

issn

volume

| | |
|---|---|
| issue | |
| pages | |
| DOI | |
| URL | |
| Source | ◯ MedRxiv<br>◯ BioRxiv<br>◯ EMBASE<br>◯ PubMed<br>◯ Other |

#### Screening

| | |
|---|---|
| Comments | |
| Help | ◯ Yes<br>◯ No |
| Reference number in publication | |
| To be included in the review | ◯ Yes ◯ No |
| Reason for exclusion | ◯ Excluded based on title/abstract<br>◯ Publication not on COVID-19<br>◯ No original data<br>◯ Data included in other publication (i.e. same population reported twice)<br>◯ Duplicate (i.e. exact same article)<br>◯ Insufficient extractable data<br>◯ Study design inappropriate<br>◯ Infections not diagnosed with a molecular test OR serologic test<br>◯ Aim of mathematical model not in review scope<br>◯ Inadequate or undocumented follow-up time<br>◯ Preprint of published article<br>◯ Other |
| Serological study | ◯ Yes<br>◯ No |
| Comment from screening | |
| Major study design group from the screening | ◯ EPI<br>◯ BASIC<br>◯ Non-Original<br>◯ NA |

| | |
|---|---|
| Major study design group | ◯ EPI<br>◯ BASIC<br>◯ Non-Original<br>◯ NA |
| Verification | |
| Verification comment | |
| Resolution comment | |

### Data extraction form

#### General Information

Record ID:

**author: [author_1] ([year]) [title]**

**General information**

Aim/objective of the study:

(As reported by author)

Study design:
◯ Case series
◯ Cross sectional study
◯ Cohort study
◯ Mathematical modelling study
◯ Case-control study
◯ Outbreak Investigation
◯ Other

Please specify "other" study design:

SARS-CoV-2 infection diagnosed by:
◯ PCR
◯ Serology
◯ Both PCR and serology

Setting of data collection:
◯ Cruise ship
◯ Hospital
◯ Traveler/evacuee from an affected area
◯ Contact tracing
◯ Health care worker
◯ Family cluster
◯ Other

Please specify "other" setting:

Setting 2 [used in Q1, Q2 analysis]
◯ 1. Contact investigation
◯ 2. Contact investigation, aggregated
◯ 3. Outbreak investigation
◯ 4. Statistical model
◯ 5. Screening
◯ 6. Hospitalised adults
◯ 7. Hospitalised children
◯ 8. Hospitalised children and adults
◯ 9. Screening: institutional setting
◯ 10. Screening: community setting
◯ 11. Screening: occupational

Country:

- ◯ Afghanistan
- ◯ Albania
- ◯ Algeria
- ◯ Andorra
- ◯ Angola
- ◯ Antigua and Barbuda
- ◯ Argentina
- ◯ Armenia
- ◯ Aruba
- ◯ Australia
- ◯ Austria
- ◯ Azawad
- ◯ Azerbaijan
- ◯ Bahamas
- ◯ Bahrain
- ◯ Bangladesh
- ◯ Barbados
- ◯ Belarus
- ◯ Belgium
- ◯ Belize
- ◯ Benin
- ◯ Bermuda
- ◯ Bhutan
- ◯ Bolivia
- ◯ Bosnia and Herzegovina
- ◯ Botswana
- ◯ Brazil
- ◯ Brunei
- ◯ Bulgaria
- ◯ Burkina Faso
- ◯ Burundi
- ◯ Cambodia
- ◯ Cameroon
- ◯ Canada
- ◯ Cape Verde
- ◯ Central African Republic
- ◯ Chad
- ◯ Chechnya
- ◯ Chile
- ◯ China
- ◯ Colombia
- ◯ Comoros
- ◯ Congo
- ◯ Costa Rica
- ◯ Côte d'Ivoire
- ◯ Croatia
- ◯ Cuba
- ◯ Curaçao
- ◯ Cyprus
- ◯ Czechoslovakia,
- ◯ Denmark
- ◯ Dominican Republic
- ◯ Ecuador
- ◯ Egypt
- ◯ El Salvador
- ◯ Equatorial Guinea
- ◯ Eritrea
- ◯ Estonia
- ◯ Ethiopia
- ◯ Fiji
- ◯ Finland
- ◯ France
- ◯ French Guyana
- ◯ Gabon
- ◯ Gambia
- ◯ Georgia
- ◯ Germany
- ◯ Ghana
- ◯ Greece

◯ Greenland
◯ Guatemala
◯ Guinea
◯ Guyana
◯ Haiti
◯ Honduras
◯ Hong Kong
◯ Hungary
◯ Iceland
◯ India
◯ Indonesia
◯ Iran
◯ Iraq
◯ Ireland
◯ Israel
◯ Italy
◯ Jamaica
◯ Japan
◯ Jordan
◯ Kazakhstan
◯ Kenya
◯ Kosovo
◯ Kuwait
◯ Kyrgyzstan
◯ Laos
◯ Latvia
◯ Lebanon
◯ Liberia
◯ Libya
◯ Liechtenstein
◯ Lithuania
◯ Luxembourg
◯ Macedonia
◯ Madagascar
◯ Malawi
◯ Malaysia
◯ Maldives
◯ Mali
◯ Malta
◯ Mauritius
◯ Mexico
◯ Moldova
◯ Monaco
◯ Mongolia
◯ Montenegro
◯ Morocco
◯ Mozambique
◯ Myanmar
◯ Multiple
◯ Namibia
◯ Nepal
◯ Netherlands
◯ New Zealand
◯ Nicaragua
◯ Niger
◯ Nigeria
◯ North Korea
◯ North Yemen
◯ Norway
◯ Oman
◯ Pakistan
◯ Palestine
◯ Panama
◯ Papua New Guinea
◯ Paraguay
◯ Peru
◯ Philippines
◯ Poland
◯ Portugal
◯ Puerto Rico
◯ Qatar

◯ Romania
◯ Russia
◯ Rwanda
◯ Samoa
◯ San Marino
◯ Saudi Arabia
◯ Senegal
◯ Serbia
◯ Seychelles
◯ Sierra Leone
◯ Singapore
◯ Slovakia
◯ Slovenia
◯ Solomon Islands
◯ Somalia
◯ South Africa
◯ South Korea
◯ South Sudan
◯ South Yemen
◯ Spain
◯ Sri Lanka
◯ Sudan
◯ Suriname
◯ Swaziland
◯ Sweden
◯ Switzerland
◯ Syria
◯ Tajikistan
◯ Tanzania
◯ Thailand
◯ Tibet
◯ Togo
◯ Tunisia
◯ Turkey
◯ Turkmenistan
◯ Uganda
◯ Ukraine
◯ United Arab Emirates
◯ United Arab Republic
◯ United Kingdom
◯ United States of America
◯ Uruguay
◯ Uzbekistan
◯ Venezuela
◯ Vietnam
◯ Yemen
◯ Yugoslavia
◯ Zambia
◯ Zanzibar
◯ Zimbabwe
◯ Sao Tome and Principe
◯ More than one country
◯ Other

---

Region or comments from "country":

---

Duration of follow-up time (choose all that apply):

☐ 14 days after the last possible exposure date
☐ 7 days after diagnosis
☐ Until negative PCR result
☐ >7 days after diagnosis

| | |
|---|---|
| Which questions are addressed in this study? | ☐ Question 1: Amongst people who become infected with SARS-CoV-2, what proportion does not experience symptoms at all during their infection?<br>☐ DO NOT CHOOSE - Question 2: Amongst people who become infected with SARS-CoV-2, what proportion has no symptoms at the time of testing, but develops symptoms later?<br>☐ Question 2.1: What is the secondary attack rate (SAR) from asymptomatic or pre-symptomatic index cases?ORQuestion 2.2: What proportion of SARS-CoV-2 infections is accounted for by people who are either asymptomatic throughout infection, or pre-symptomatic? |
| Comments: | (Enter "none" if no comments) |
| Extracted by: | (Last name only) |
| Verified by: | (Last name only) |

### Question 1

Record ID:

**author: [author_1] ([year]) [title]**

**Question 1: Amongst people who become infected with SARS-CoV-2, what proportion does not experience symptoms at all during their infection?**

**Reminder: Question 1: Proportion or rate of people with asymptomatic SARS-CoV-2 infections who do not experience symptoms at all during their infection (i.e. from the time of testing to the end of the follow-up);**

| | |
|---|---|
| Number of clusters described: | ◯ 1<br>◯ 2<br>◯ 3<br>◯ 4<br>◯ NA |
| Omit from meta-analysis | ◯ Yes<br>◯ No |
| Number of individuals that were asymptomatic throughout infection: | |
| Total number of individuals (denominator): | |
| Number of individuals that were asymptomatic throughout infection [cluster2]: | |
| Total number of individuals (denominator) [cluster2]: | |
| Number of individuals that were asymptomatic throughout infection [cluster3]: | |
| Total number of individuals (denominator) [cluster3]: | |

**Follow-up time**

Please describe the follow-up time in days. The description should include how follow-up times were measured and for which people follow-up times were reported.

#### Sex

Number of females who remained asymptomatic throughout infection:

(Enter "9999" if not reported.)

Number of males who remained asymptomatic throughout infection:

(Enter "9999" if not reported.)

#### Age

Enter median age of all people who remained asymptomatic throughout infection:

(Enter "9999" if not reported.)

Enter IQR for age of all people who remained asymptomatic throughout infection:

(Enter "9999" if not reported.)

Age range of asymptomatic

Select one or more if applicable

- ☐ Children (< 18 years)
- ☐ Adults (18 - 65 years)
- ☐ Older adults (>65 years)
- ☐ All ages
- ☐ Not Reported

#### Comments

Comments:

Extracted by:

(Enter last name)

Verified by:

(Enter last name)

### Question 2.1 and 2.2

Record ID:

**author: [author_1] ([year]) [title]**

What does this study report?

◯ Q. 2.1 SAR
◯ Q. 2.2 Transmission proportion
◯ Both
◯ NA

Summary of results:

**Question 2.1**
**What is the secondary attack rate from asymptomatic or pre-symptomatic index cases?**

**SAR (Secondary attack rate)**
**Number of infections (numerator) caused by certain group of people: Here asymptomatics vs symptomatics (denominator)**

Asymptomatic SAR

(number infected;number asymptomatic)

Asymptomatic SAR Risk Ratio (RR)

(Enter "9999" if not reported.)

Asymptomatic SAR Risk Ratio (RR) Lower Confidence Interval

(Enter "9999" if not reported.)

Asymptomatic SAR Risk Ratio (RR) Upper Confidence Interval

(Enter "9999" if not reported.)

Asymptomatic SAR Odds Ratio (OR)

(Enter "9999" if not reported.)

Asymptomatic SAR Odds Ratio (OR) Lower Confidence Interval

(Enter "9999" if not reported.)

Asymptomatic SAR Odds Ratio (OR) Upper Confidence Interval

(Enter "9999" if not reported.)

Pre-symptomatic SAR

(number infected;number pre-symptomatic)

| | |
|---|---|
| Pre-symptomatic SAR Risk Ratio (RR) | (Enter "9999" if not reported.) |
| Pre-symptomatic SAR Risk Ratio (RR) Lower Confidence Interval | (Enter "9999" if not reported.) |
| Pre-symptomatic SAR Risk Ratio (RR) Upper Confidence Interval | (Enter "9999" if not reported.) |
| Pre-symptomatic SAR Odds Ratio (OR) | (Enter "9999" if not reported.) |
| Pre-symptomatic SAR Odds Ratio (RR) Lower Confidence Interval | (Enter "9999" if not reported.) |
| Pre-symptomatic SAR Odds Ratio (OR) Upper Confidence Interval | (Enter "9999" if not reported.) |
| Mild symptomatic SAR | (number infected;number mild symptomatic) |
| Moderate symptomatic SAR | (number infected;number mild symptomatic) |
| Severe/critical symptomatic SAR | (number infected;number critical symptomatic) |
| ANY symptomatic SAR | (number infected;number ANY symptomatic) |

**Question 2.2: What proportion of SARS-CoV-2 infections is accounted for by people who are either asymptomatic throughout infection, or pre-symptomatic?**

| | |
|---|---|
| Region/setting | (If multiple settings are described, please specify and separate with ';') |

#### Asymptomatic transmission

Proportion asymptomatic transmission (median)

(For all fields: if mutiple settings are described use ';' to separate extraction)

Proportion asymptomatic transmission (lower CrI)

Proportion asymptomatic transmission (upper CrI)

#### Pre-symptomatic transmission

Proportion pre-symptomatic transmission (median)

(For all fields: if mutiple settings are described use ';' to separate extraction)

Proportion pre-symptomatic transmission (lower CrI)

Proportion pre-symptomatic transmission (upper CrI)

Comment:

#### Extraction

Extracted by:

(Enter last name)

Verfied by:

(Enter last name)

### Risk of Bias - observational EPI [update 3]

Record ID:

ROB assigned to [rob reviewer 1]; performed by:

((e.g., NL))

ROB assigned to [rob reviewer 2]; performed by:

( (e.g., NL))

Resolution assigned to [rob reviewer 3]; performed by:

(R1) If the study reports more than one cluster, please describe the participants of cluster 1 and cluster 2:

(R2) If the study reports more than one cluster, please describe the participants of cluster 1 and cluster 2:

#### Selection Bias

(R1) Reporting: How was the target population described?

((Extract it from the study))

(R1) Reporting: How was the target population described in cluster 2?

((Extract it from the study))

(R2) Reporting: How was the target population described?

((Extract it from the study))

(R2) Reporting: How was the target population described in cluster 2?

((Extract it from the study))

(R1) Question 1- Was the sample invited to participate a close or true representation of the target population?

◯ High ◯ Unclear ◯ Low

(R1) Question 1- Was the sample invited in cluster 2 to participate a close or true representation of the target population?

◯ High ◯ Unclear ◯ Low
◯ NA

(R1) Comments

| | |
|---|---|
| (R2) Question 1- Was the sample invited to participate a close or true representation of the target population? | ◯ High ◯ Unclear ◯ Low |
| (R2) Question 1- Was the sample invited in cluster 2 to participate a close or true representation of the target population? | ◯ High ◯ Unclear ◯ Low ◯ NA |
| (R2) Comments | |
| (Consensus) Question 1- Was the sample invited to participate a close or true representation of the target population? | ◯ High ◯ Unclear ◯ Low |
| (Consensus) Question 1- Was the sample invited in cluster 2 to participate a close or true representation of the target population? | ◯ High ◯ Unclear ◯ Low |

#### Response Rate

| | |
|---|---|
| (R1) Reporting: Was the response rate from the eligible population provided? | ◯ Yes ◯ No |
| (R1) Reporting: Was the response rate from the eligible population in cluster 2 provided? | ◯ Yes ◯ No ◯ NA |
| (R2) Reporting: Was the response rate from the eligible population provided? | ◯ Yes ◯ No |
| (R2) Reporting: Was the response rate from the eligible population in cluster 2 provided? | ◯ Yes<br>◯ No<br>◯ NA |
| (R1) If the response rate is not provided. Can you calculate the response rate? | ◯ Yes ◯ No ◯ NA |
| (R1) If the response rate for cluster 2 is not provided. Can you calculate the response rate for cluster 2? | ◯ Yes ◯ No ◯ NA |
| (R2) If the response rate is not provided. Can you calculate the response rate? | ◯ Yes ◯ No ◯ NA |
| (R2) If the response rate for cluster 2 is not provided. Can you calculate the response rate for cluster 2? | ◯ Yes<br>◯ No<br>◯ NA |
| (R1) Number of individuals who participated in the study | |
| (R1) Number of individuals in cluster 2 who participated in the study | |
| (R2) Number of individuals who participated in the study | |

(R2) Number of individuals in cluster 2 who participated in the study

(R1) Number of eligible participants

(R1) Number of eligible participants for cluster 2

(R2) Number of eligible participants

(R2) Number of eligible participants for cluster 2

(R1) Response rate (%)

(R1) Response rate (%) for cluster 2

(R2) Response rate (%)

(R2) Response rate (%) for cluster 2

(R1) Comments

(R2) Comments

(Consensus) Response rate (%)

(Consensus) Response rate (%) for cluster 2

#### Selection Bias - Response Rate

(R1) Question 2- The characteristics of non-respondents, if any, who were eligible are similar to those who participated in the study?

◯ High ◯ Unclear ◯ Low

(R1) Question 2- The characteristics of non-respondents for cluster 2, if any, who were eligible are similar to those who participated in the study?

◯ High
◯ Unclear
◯ Low
◯ NA

(R1) Comments

| | |
|---|---|
| (R2) Question 2- The characteristics of non-respondents, if any, who were eligible are similar to those who participated in the study? | ◯ High ◯ Unclear ◯ Low |
| (R2) Question 2- The characteristics of non-respondents, if any, who were eligible are similar to those who participated in the study for cluster 2? | ◯ High<br>◯ Unclear<br>◯ Low<br>◯ NA |
| (R2) Comments | |
| (Consensus) Q2- The characteristics of non-respondents, if any, who were eligible are similar to those who participated in the study? | ◯ High ◯ Unclear ◯ Low |
| (Consensus) Q2- The characteristics of non-respondents, if any, who were eligible are similar to those who participated in the study for cluster 2? | ◯ High ◯ Unclear ◯ Low |

#### Information Bias

| | |
|---|---|
| (R1) Reporting: How was symptomatic and asymptomatic defined? | |
| (R2) Reporting: How was symptomatic and asymptomatic defined? | |
| (R1) Question 3 - Was the assessment of symptoms status adequate? | ◯ High ◯ Unclear ◯ Low |
| (R1) Question 3 - Was the assessment of symptoms status adequate for cluster 2? | ◯ High<br>◯ Unclear<br>◯ Low<br>◯ NA |
| (R1) Comments | |
| (R2) Question 3 - Was the assessment of symptoms status adequate ? | ◯ High ◯ Unclear ◯ Low |
| (R2) Question 3 - Was the assessment of symptoms status adequate for cluster 2? | ◯ High<br>◯ Unclear<br>◯ Low<br>◯ NA |
| (R2) Comments | |
| (Consensus) Q3- Was the assessment of symptoms status adequate? | ◯ High ◯ Unclear ◯ Low |
| (Consensus) Q3- Was the assessment of symptoms status adequate for cluster 2? | ◯ High ◯ Unclear ◯ Low |

| | |
|---|---|
| (R1) Question 4- Based on the method symptoms were collected, is there a risk of recall bias? | ○ High ○ Unclear ○ Low |
| (R1) Question 4- Based on the method symptoms were collected, is there a risk of recall bias for cluster 2? | ○ High ○ Unclear ○ Low ○ NA |
| (R1) Comments | |
| (R2) Question 4- Based on the method symptoms were collected, is there a risk of recall bias? | ○ High ○ Unclear ○ Low |
| (R2) Question 4- Based on the method symptoms were collected, is there a risk of recall bias for cluster 2? | ○ High ○ Unclear ○ Low ○ NA |
| (R2) Comments | |
| (Consensus) Q4- Based on the method symptoms were collected, is there a risk of recall bias? | ○ High ○ Unclear ○ Low |
| (Consensus) Q4- Based on the method symptoms were collected, is there a risk of recall bias for cluster 2? | ○ High ○ Unclear ○ Low |
| (R1) Reporting: For how long were the participants followed to assess symptom status? | ☐ 14 days after the last possible exposure date ☐ 7 days after diagnosis<br>☐ Until negative PCR result<br>☐ Both 14 days after the last possible exposure date AND 7 days after diagnosis<br>☐ Longer follow-up |
| (R1) Reporting: For how long were the participants followed to assess symptom status in cluster 2? | ☐ 14 days after the last possible exposure date<br>☐ 7 days after diagnosis<br>☐ Until negative PCR result<br>☐ Both 14 days after the last possible exposure date AND 7 days after diagnosis<br>☐ Longer follow-up<br>☐ NA |
| (R2) Reporting: For how long were the participants followed to assess symptom status? | ☐ 14 days after the last possible exposure date ☐ 7 days after diagnosis<br>☐ Until negative PCR result<br>☐ Both 14 days after the last possible exposure date AND 7 days after diagnosis<br>☐ Longer follow-up |
| (R2) Reporting: For how long were the participants followed to assess symptom status? | ☐ 14 days after the last possible exposure date<br>☐ 7 days after diagnosis<br>☐ Until negative PCR result<br>☐ Both 14 days after the last possible exposure date AND 7 days after diagnosis<br>☐ Longer follow-up<br>☐ NA |

| | |
|---|---|
| (Consensus) Reporting: For how long were the participants followed to assess symptom status? | ☐ 14 days after the last possible exposure date ☐ 7 days after diagnosis<br>☐ Until negative PCR result<br>☐ Both 14 days after the last possible exposure date AND 7 days after diagnosis<br>☐ Longer follow-up |
| (Consensus) Reporting: For how long were the participants followed to assess symptom status? | ○ High ○ Unclear ○ Low |
| (R1) Reporting: Was the viral load reported? Please provide information about it CT (cycle threshold) values. | (If not reported, please write 'NR') |
| (R2) Reporting: Was the viral load reported? Please provide information about it CT (cycle threshold) values. | (If not reported, please write 'NR') |
| (R1) Question 5- Is there a risk that asymptomatic status was misclassified? | ○ High ○ Unclear ○ Low |
| (R1) Question 5- Is there a risk that asymptomatic status was misclassified in cluster 2? | ○ High ○ Unclear ○ Low<br>○ NA |
| (R1) Comments | |
| (R2) Question 5- Is there a risk that asymptomatic status was misclassified? | ○ High ○ Unclear ○ Low |
| (R2) Question 5- Is there a risk that asymptomatic status was misclassified in cluster 2? | ○ High ○ Unclear ○ Low<br>○ NA |
| (R2) Comments | |
| (Consensus) Q5- Is there a risk that asymptomatic status was misclassified? | ○ High ○ Unclear ○ Low |
| (Consensus) Q5- Is there a risk that asymptomatic status was misclassified in cluster 2? | ○ High ○ Unclear ○ Low |

**Selective or Incomplete Reporting of Outcome**

| | |
|---|---|
| (R1) Reporting: The authors present the symptom status of all participants at the end of follow-up? | ○ Yes ○ No |
| (R1) Reporting: The authors present the symptom status of all participants in cluster 2 at the end of follow-up? | ○ Yes ○ No ○ NA |
| (R2) Reporting: The authors present the symptom status of all participants at the end of follow-up? | ○ Yes ○ No |

| | |
|---|---|
| (R2) Reporting: The authors present the symptom status of all participants in cluster 2 at the end of follow-up? | ◯ Yes ◯ No ◯ NA |
| (R1) Question 6-Is there a risk of incomplete or selective reporting of symptoms status among those who were positive for SARS-CoV-2? | ◯ High ◯ Unclear ◯ Low |
| (R1) Question 6-Is there a risk of incomplete or selective reporting of symptoms status among those who were positive for SARS-CoV-2 in cluster 2? | ◯ High ◯ Unclear ◯ Low ◯ NA |
| (R1) Comments | |
| (R2) Question 6-Is there a risk of incomplete or selective reporting of symptoms status among those who were positive for SARS-CoV-2? | ◯ High ◯ Unclear ◯ Low |
| (R2) Question 6-Is there a risk of incomplete or selective reporting of symptoms status among those who were positive for SARS-CoV-2 in cluster 2? | ◯ High ◯ Unclear ◯ Low ◯ NA |
| (R2) Comments | |
| (Consensus) Q6- Is there a risk of incomplete or selective reporting of symptoms status? | ◯ High ◯ Unclear ◯ Low |
| (Consensus) Q6- Is there a risk of incomplete or selective reporting of symptoms status in cluster 2? | ◯ High ◯ Unclear ◯ Low |

**Final Comments**

| | |
|---|---|
| (R1) Is there a risk of over or under-estimating the proportion of the truly asymptomatic population? | ◯ Underestimation of the proportion of asymptomatics ◯ Overestimation of the proportion of asymptomatics ◯ The proportion provided by the authors is accurate ◯ Unclear |
| (R1) Is there a risk of over or under-estimating the proportion of the truly asymptomatic population in cluster 2? | ◯ Underestimation of the proportion of asymptomatics ◯ Overestimation of the proportion of asymptomatics ◯ The proportion provided by the authors is accurate ◯ Unclear ◯ NA |
| (R1) Comments | |
| (R2) Is there a risk of over or under-estimating the proportion of the truly asymptomatic population? | ◯ Underestimation of the proportion of asymptomatics ◯ Overestimation of the proportion of asymptomatics ◯ The proportion provided by the authors is accurate ◯ Unclear |

| | |
|---|---|
| (R2) Is there a risk of over or under-estimating the proportion of the truly asymptomatic population in cluster 2? | ◯ Underestimation of the proportion of asymptomatics ◯ Overestimation of the proportion of asymptomatics ◯ The proportion provided by the authors is accurate ◯ Unclear ◯ NA |
| (R2) Comments | |
| Reviewer 3 (Consensus) - comments | |

#### S2 Appendix. Analysis of other systematic reviews of asymptomatic SARS-CoV-2 infection

| First author, publication year [ref] | Studies included (n) | Search dates | Inclusion criteria[a] | Summary estimate[b] (95% CI) | $I^2$ | $\tau^2$ | Prediction interval |
|---|---|---|---|---|---|---|---|
| Byambasuren, 2020 [1] | 13 | Until 20 July 2020 | "primary studies on asymptomatic prevalence in which (1) the sample frame includes at-risk populations and (2) follow-up was sufficient to identify pre-symptomatic cases" | 18% (11% - 27%) | 81% | 0.6746 | 3% - 59% |
| Beale, 2020 [2] | 21 | Until 25 August 2020 | "studies based in community settings that involved systematic PCR testing on participants and follow-up symptom monitoring regardless of symptom status." | 21% (13% - 33%) | 86% | 1.5845 | 2% - 80% |
| Chen 2021 [3] | 241 | Until 31 December 2020 | "Original investigations with sample size (or number of subjects) not less than five were included for further analyses." | 20% (17% - 23%) | 99% | 2.3483 | 1% - 83% |
| Ma 2021 [4] | 95 | Until 4 February 2021 | "Cross-sectional studies, cohort studies, case series studies, and case series on transmission reporting the number of asymptomatic infections among the tested and confirmed COVID-19 populations that were published in Chinese or English were included." | 51% (42% - 60%) | 97% | 2.7950 | 4% - 97% |
| | 77[c] | | | 38% (32% - 44%) | 97% | 1.2619 | 6% - 85% |
| Sah 2021 [5] | 170 | Until 2 April 2021 | "Studies that reported silent infections at the time of testing, whether presymptomatic or asymptomatic. Index cases were removed to minimize representational bias that would result in overestimation of symptomaticity." | 36% (31% - 41%) | 94% | 1.8989 | 3% - 89% |

[a]Text extracted from study abstract.
[b]Using the random effects model and the logit transformation to calculate a summary estimate in the *metaprop* package in R. All studies published summary estimates, 95% CI, and $I^2$ values using their own methodology. Only one study (Beale) published a $\tau^2$ value. None of the studies published a prediction interval.
[c]Excluding studies in which all cases were asymptomatic.

**Figure. Graphical representation of summary estimates of asymptomatic SARS-CoV-2 infection extracted from other systematic reviews**

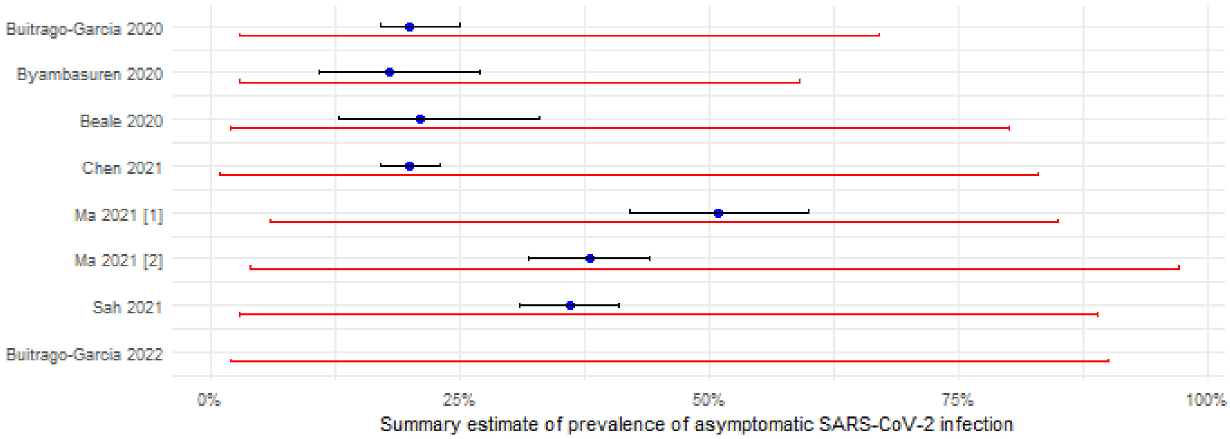

Summary estimate as reported by authors is blue with black lines indicating the 95% confidence interval.
Prediction interval, calculated from study data, represented as red line.

[1] Including all studies
[2] Excluding studies in which all cases were asymptomatic.
